# Sex Differences in Primary Care Contacts and Recurrent Overdose Risk in British Columbia

**DOI:** 10.64898/2026.10.09.26365159

**Authors:** Jennifer Vincent, Heather Palis

## Abstract

**Background:** Risk of recurrent overdose is elevated following non-fatal overdose (NFOD) survival; timely interventions may reduce this risk. The specific role of mental health care in reducing recurrence is relatively underexplored, and post-overdose care has seldom been examined for sex differences. This study examined sex differences in the association between mental health-related primary care contacts following NFOD and overdose recurrence risk among people in British Columbia (BC) with a documented depression diagnosis history.

**Methods:** This was a nested case-control study using linked administrative health data from the Provincial Health Services Authority’s Platform for Analytics and Data. The source cohort (n=7,022) comprised BC residents with a history of a documented depression diagnosis between 2010-2014 and at least one NFOD between 2015-2024. Cases experienced a recurrent overdose within one year of their index overdose; controls were selected using incidence density sampling. Exposures were the presence and number of mental health related primary care contacts in the 30-or 90-day window following the index overdose. Conditional logistic regression models were used to estimate adjusted incidence rate ratios (aIRRs) and confidence intervals (CI), overall and by sex. Secondary analyses were conducted using psychiatrist contacts.

**Results:** Mental-health related primary care contact rates were low, with only 16.3% and 28.1% of the cohort with any contact within 30 and 90 days, respectively. Mental health-related primary care contacts were more strongly associated with reduced recurrence risk in the 30-day window (any contact aIRR: 0.57 [CI: 0.46-0.71]) than in the 90-day window (aIRR: 0.78 [CI: 0.65-0.94]). Any contact within 30 days was more strongly associated with lower recurrence risk among males (aIRR: 0.53 [CI: 0.39-0.72]) than among females (aIRR: 0.67 [CI: 0.50-0.91]), whereas associations between the number of contacts and recurrence risk were similar for males and females. Any psychiatrist contact within 30 days was more strongly associated with lower recurrence risk among females (aIRR: 0.50 [CI: 0.34-0.73]) than males (aIRR: 0.70 [0.52-0.95]).

**Discussion:** Mental health-related care contacts shortly following NFOD were associated with lower overdose recurrence risk among people with a documented depression diagnosis history, with modest sex differences that varied by provider type. Low rates of contacts overall suggest gaps in access to care following NFOD, particularly among males. Further research on sex differences in post-overdose care are warranted.

## Background

In the weeks to months following a non-fatal overdose, there is a risk of recurrent overdose, particularly within the first month, with risks persisting up to a year ^1–6^. Prior work suggests that timely interventions during high-risk periods help reduce recurrent overdose risk ^5,7^. Substance use and harm reduction interventions, such as opioid agonist treatment, naloxone distribution, and behavioural interventions, have been found to be protective against overdose recurrence in this period ^6–9^.

Depression has previously been identified as a risk factor for overdose recurrence ^6^; however, existing literature has more commonly on substance use interventions, leaving the specific role of mental health care in reducing recurrence relatively underexplored ^7,9^. Unmet mental health needs have been reported by those who have experienced an overdose ^10^, and not all people who experience an overdose have substance use disorder ^11^; these people may have other needs to address to prevent recurrence. For these people, identifying interventions to reduce risk of recurrent overdose is particularly important.

Evidence from substance use and mental health treatment more broadly suggests sex and gender differences in treatment access, responses to treatments, and types of treatments initiated ^12–15^. Some research indicates that addressing mental health needs may be of particular benefit for women in substance use treatment ^13^, and unmet mental health needs may have greater impacts on overdose risk among women and females compared to men and males ^16–18^. Yet post-overdose interventions have seldom been examined for sex and/or gender differences.

### Gap, Objective, and Hypothesis

#### Gap

Despite the importance of timely post-overdose care in reducing recurrent overdose risk, few studies on post-overdose health service contacts, broadly and for mental health specifically, disaggregate outcomes by sex or gender. This gap is particularly important given the evidence that people have reported unmet mental health needs following a non-fatal overdose, and that unmet mental health needs are associated with overdose risk, with potentially stronger associations among women ^16–18^.

#### Objective

To examine sex-differences in the association between mental-health related primary care contact following first overdose and risk of recurrent overdose among people with a documented history of a depression diagnosis.

#### Hypothesis

The association between mental health related primary care contacts and overdose risk is greater for females than for males.

## Methods

### Study Design

This was a nested case-control study using linked administrative health data in British Columbia, Canada. The source cohort (N = 7,022) comprised people with a documented depression diagnosis history and at least one non-fatal overdose identified during the study period of January 1, 2015, to December 31, 2024. Exposures of mental health related primary care contacts, measured in the 30-or 90-day period following a person’s index overdose, were examined for associations with recurrent overdose within one year of the index overdose. The study period spanned January 1, 2015, to December 31, 2024.

### Data sources

Data for this study was drawn from the Provincial Health Services Authority’s Platform for Analytics and Data (PANDA). PANDA contains linked administrative health data for all residents in British Columbia. These datasets are linked at the individual level using the PANDA Patient Matching Service algorithm which first applies deterministic matching using Personal Health Numbers, followed by a series of deterministic and non-deterministic matches by sex, name, and date of birth ^19^. This platform and its linkages evolved from a proof-of-concept developed during the acute phase of the COVID-19 pandemic, as described by Wilton et al. ^20,21^.

The following datasets were used in this analysis: Medical Services Plan (MSP), Discharge Abstract Database (DAD), BC Emergency Health Services (BCEHS), BC Coroners Service (BCCS), and Client Roster (CLR)^22–26^. In addition, data were derived from the National Ambulatory Care Reporting System (NACRS) and the Institut national de santé publique du Québec (INSPQ) (See Table 1 for full description)^27,28^.

**Table 1.** Datasets Used in the Analysis of Recurrent Overdose Risk among People with a History of a Depression Diagnosis, British Columbia, 2015-2024.

| *Dataset* | *Use* |
| --- | --- |
| National Ambulatory Care Reporting System (NACRS) | Used to identify people with no fixed address, in conjunction with DAD. |
| Institut national de santé publique du Québec (INSPQ) | Used to derive area-based measures of material and social deprivation, linked to client locations via the CLR. |
| Client Roster (CLR) | Used to derive client's age, sex at birth, and loss of follow up due to death from non-overdose causes or departure from BC. |
| Medical Services Plan (MSP) | Used to identify outpatient physician visits for ascertainment of depression and/or SUD diagnoses, in conjunction with DAD. |
| Discharge Abstract Database (DAD) | Used to identify hospital records for ascertainment of depression and/or SUD diagnoses, in conjunction with MSP. |
| BC Emergency Health Services (BC EHS) | Used to identify NFOD cases, using paramedic attended overdose records. |
| BC Coroners Service (BCCS) | Used to identify FOD cases, based records of deaths due to overdose. |

*SUD = substance use disorder; NFOD = non-fatal overdose; FOD = fatal overdose.*
*CLR, MSP, and DAD data are sourced from the BC Ministry of Health; BCCS data are sourced from BC Coroners Service; BC EHS data are sourced from the Provincial Health Services Authority.*
*NACRS data are sourced from the BC Ministry of Health; INSPQ data are sourced from the Institut national de santé publique du Québec.*

Ethics approval for this study was provided by the UBC REB under approval H25-01385. All inferences, opinions, and conclusions drawn in this publication are those of the author(s), and do not reflect the opinions or policies of the Data Steward(s), including the Provincial Health Services Authority (PHSA), Ministry of Health (MoH), BC Emergency Health Services (BCEHS), and BC Coroners Service (BCCS).

### Study Sample

The source cohort consisted of BC residents with a documented depression diagnosis between January 1, 2010, and December 31, 2014, who subsequently experienced at least one non-fatal overdose between January 1, 2015, and December 31, 2024. Follow-up time was limited to one year following each cohort member’s index overdose, defined as their first recorded non-fatal overdose during the study period. Cohort members whose index overdose occurred within the year preceding the study end (December 31, 2024) would not have had a full year of follow-up available and were included in the analysis, but follow-up time for these people was censored at study end.

Cases were defined as individuals who experienced a second overdose, either fatal or non-fatal, within one year of their index overdose, consistent with prior research on the period of elevated recurrent overdose risk following first overdose ^6^. To ensure that the recurrent overdose occurred after the exposure measurement window, a washout period was applied: for analyses with a 30-day exposure window, cases experiencing a recurrent overdose within 30 days of their index overdose were excluded; or analyses with a 90-day window, cases experiencing a recurrent overdose within 90 days of their index overdose were excluded. Figures 1 and 2 depict the study sample flow and resulting sample sizes.

**Figure 1.**
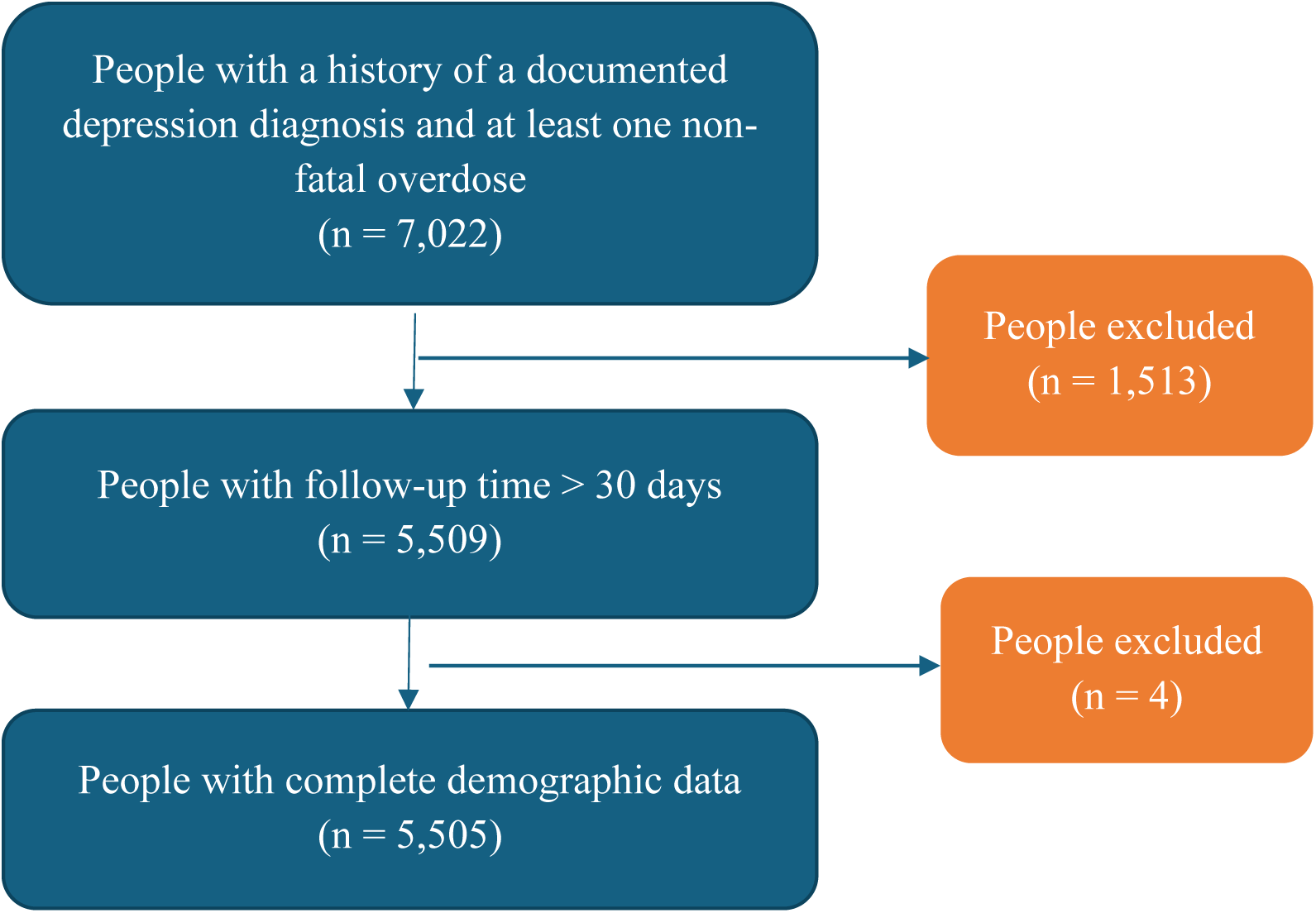
Study Sample Flow for Analyses Using the 90-day Exposure Window Among People with a History of a Documented Depression Diagnosis and a Non-Fatal Overdose, 2015-2024.

**Figure 2.**
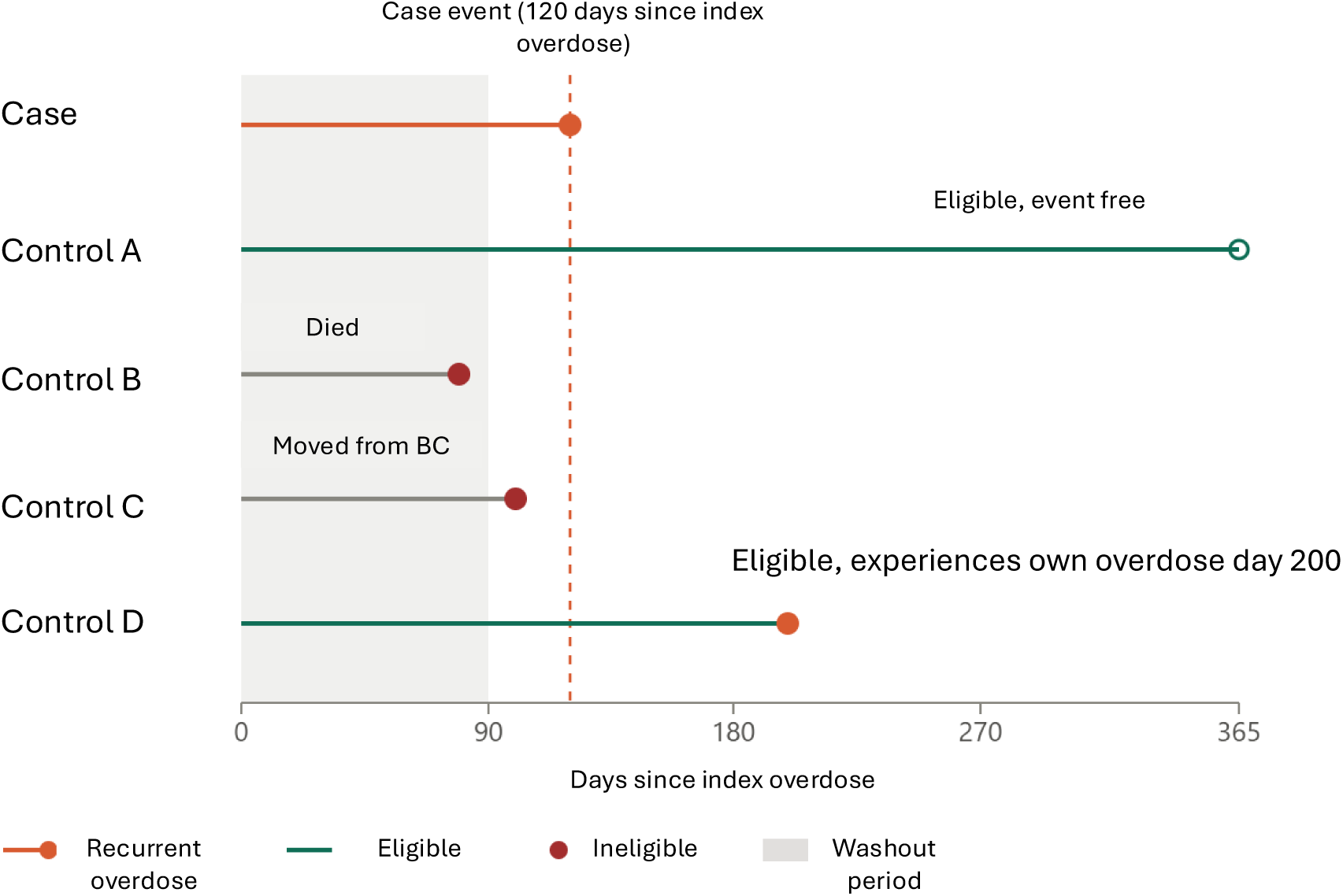
Illustration of Incidence Density Sampling with 1:n Control Selection Matched on Elapsed Time Since Index Overdose.

Controls were selected using 1:n incidence density sampling. At the elapsed time since a case’s index overdose that they experienced a recurrent overdose, cohort members were evaluated for eligibility as controls at the same elapsed time since their index overdose. Cohort members were eligible if, at that elapsed time: they had not experienced a recurrent overdose; had not died of a non-overdose cause, as determined by CLR data; had not moved out of BC, as determined by CLR data; and, had not reached the end of the study period (December 31, 2024). Individuals could serve as controls for more than one case, provided they were still eligible controls at the relevant time point. Follow-up began after the washout period of either 30 or 90 days since the index overdose, depending on the exposure variable being tested, and ended at one year since the index overdose. Individuals who were hospitalized for the entirety of either the 30 or 90 day exposure windows were included in the analysis; these accounted for approximately 1% of the study sample. For a diagram illustrating this sampling methodology, see Figure 3. The 1:n sampling approach was chosen to maximize statistical efficiency given the available data, and because prior literature suggests that increasing the number of controls relative to cases can improve precision when the distribution of the exposure is skewed ^29^. This led to a mean of 4,408 controls per case for analyses using the 30-day window, and a mean of 4,193 controls per case for analyses using the 90-day window.

**Figure 3.**
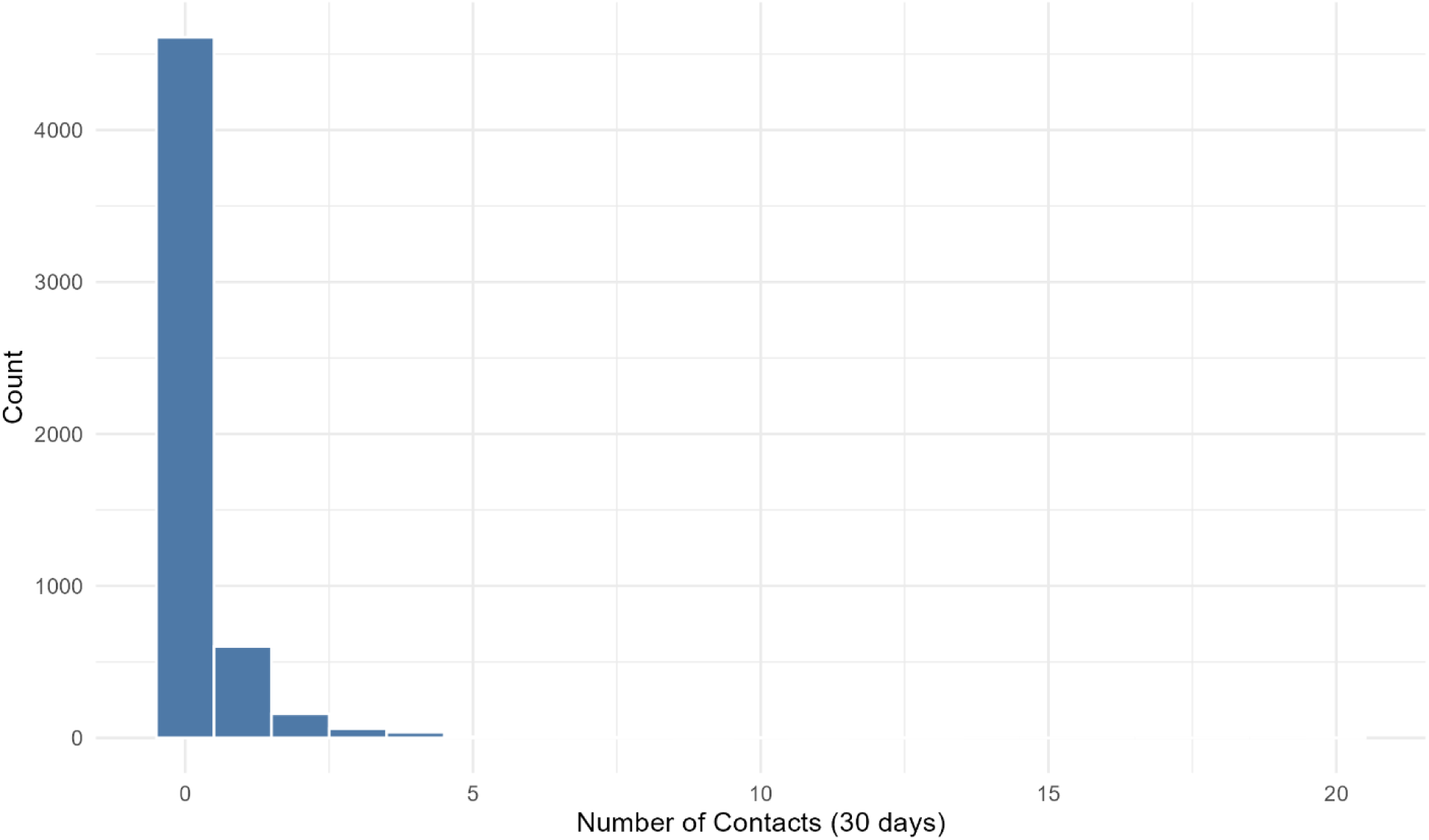
Distribution of the Number of Mental Health Related Primary Care Contacts within 30 Days of a Non-fatal Overdose, Among People with a History of a Documented Depression Diagnoses, British Columbia, 2015-2024.

**Figure 4.**
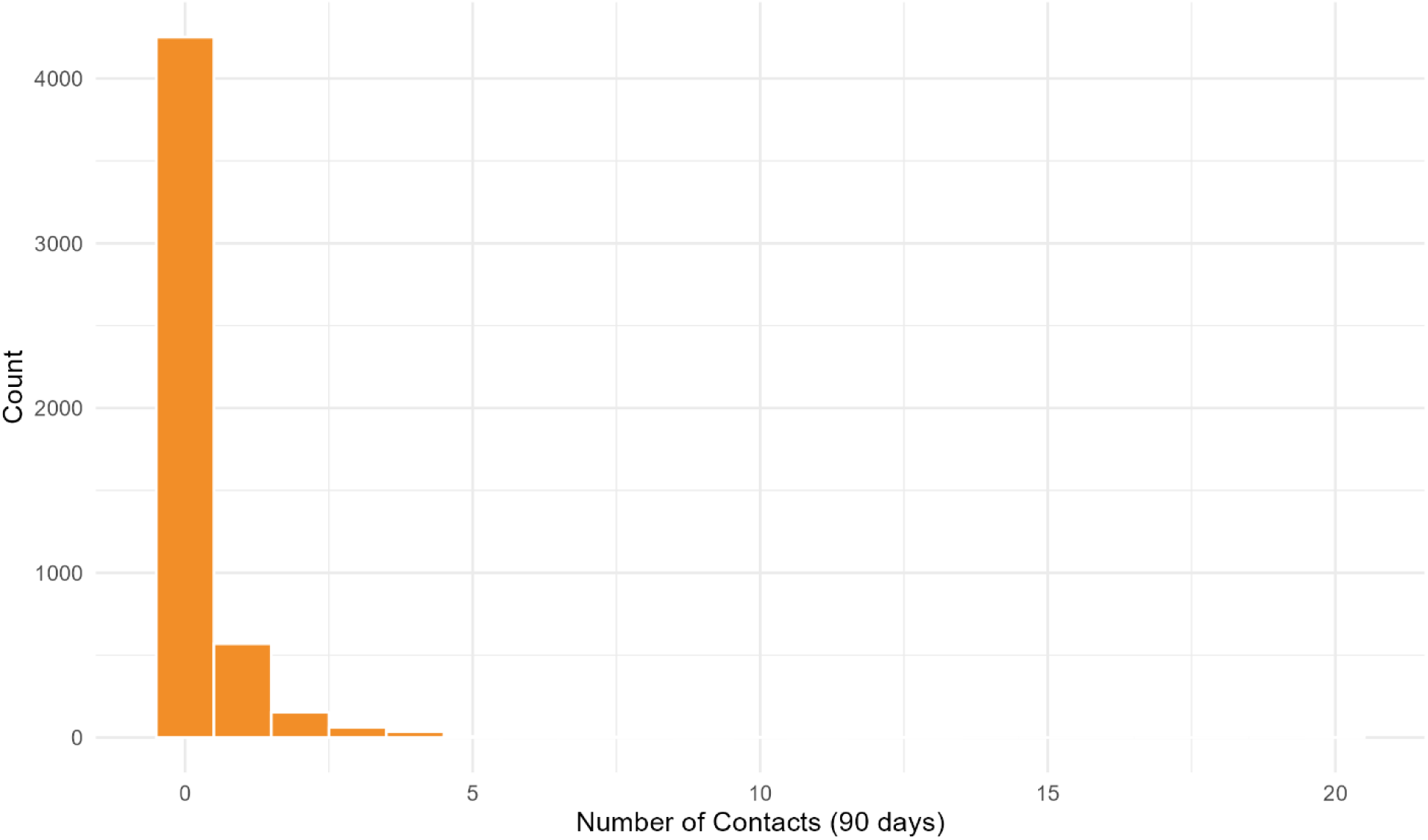
Distribution of the Number of Mental Health Related Primary Care Contacts within 90 Days of a Non-fatal Overdose among People with a History of a Documented Depression Diagnoses, British Columbia, 2015-2024.

**Figure 5.**
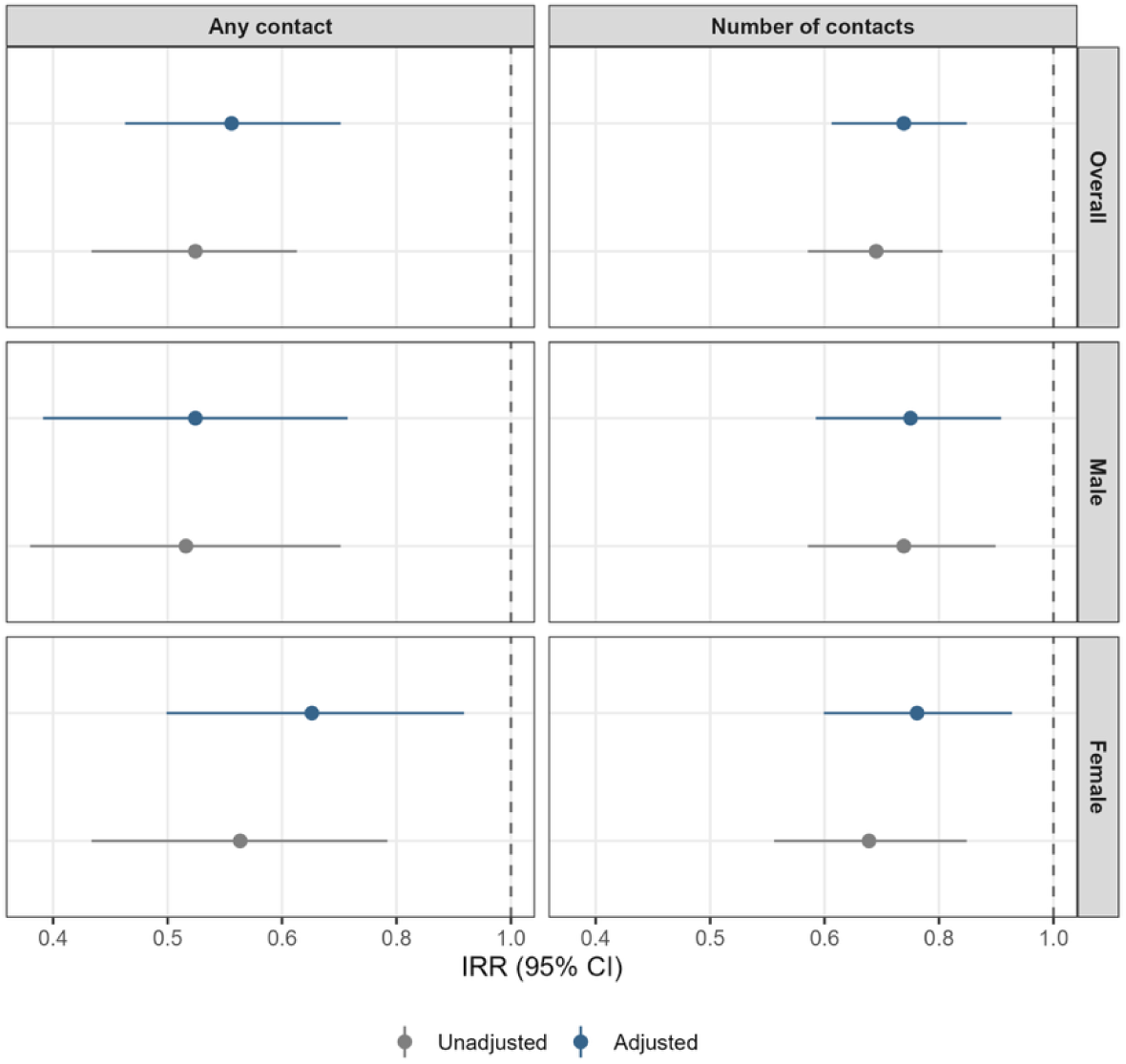
Forest Plot of Incidence Rate Ratios and 95% Confidence Intervals for Mental Health-Related Primary Care Contacts within 30-days Post Overdose and Rates of Recurrent Overdose Among People with a History of a Depression Diagnosis.

### Variables

#### Exposure

##### Primary care contacts for mental health exposure

Primary care contacts for mental health were used to create four exposure variables: the presence of a mental health related primary care contact within 30 days, the number of mental health related primary care contacts within 30 days, the presence of a mental health related primary care contact within 90 days, and the number of mental health related primary care contacts within 90 days. Using the MSP dataset, physician contacts where the billing specialty was listed as a general practitioner, and where ICD-9 codes in any diagnostic position for the contact aligned with the codes used for ascertaining mental health conditions (depression, mood and anxiety, schizophrenia and delusional disorder) within the Chronic Disease Registry ^30^ were identified as mental health related primary care contacts (Table 2).

**Table 2.** ICD-9 and 10 Codes Used for Ascertainment of Mental Health Related Care Contacts within 30 and 90 days of a Non-Fatal Overdose.

| Diagnosis | ICD-9 Codes |
| --- | --- |
| Depression | 296<br>311<br>50B (MSP only) |
| Mood/Anxiety Disorders | 296<br>300<br>311<br>50B |
| Schizophrenia and Delusional Disorders | 295 |

*MSP = Medical Services Plan. ICD = International Disease Classification. ICD-9 and ICD-10 are diagnostic coding systems used to document and classify diseases, health conditions, and healthcare encounters in a standardized fashion. ICD-9 code 50B is a BC-specific code used in MSP billing for anxiety and depression.*

For each cohort member, the number of primary care contacts for mental health within the exposure window of either 30 or 90 days was summed to determine the total number of contacts in each window. A binary variable was also created for each exposure window by indicating whether the individual had at least one mental health related primary care contact within that window.

#### Outcome

Recurrent non-fatal overdoses (NFOD) and fatal overdoses (FOD) were ascertained using BCCS and BCEHS datasets, using case-finding algorithms previously documented^31,32^. Where multiple recurrent overdoses occurred within the follow-up window, only the overdose occurring closest in time to the index overdose was selected.

#### Covariates

Covariate selection considered individual (sex, age, No Fixed Address) and interpersonal (Health Authority, Material and Social Deprivation) level factors based on known risk factors for overdose and depression and was guided by Latimore’s Social Ecological model of substance use ^33^.

##### Sex

Sex was defined as sex at birth, as recorded in the CLR and measured at the start of the study period.

##### Age group

Age was derived from the birth year and month in the CLR, in which the date of birth is set to the first day of the month in which the person was born, to protect privacy. Age in years was calculated as the difference between this date and the study start date, which was then classified into the following groups: 19 -29, 30-39, 40-49, 50-59, 60+.

##### No Fixed Address

A binary variable indicating whether a cohort member had a history of having no fixed address was derived from DAD and NACRS, using the presence of a contact listing ICD code Z-59 and forwarding station address of XX. This was derived based on the available data five-years prior to study start (January 1, 2015).

##### Health Authority

Health Authority was determined using the client’s listed community health service delivery area in the Client Roster at study start (January 1, 2015).

##### Material and Social Deprivation

Material and social deprivation, in quintiles, was derived from the Institut national de santé publique du Québec (INSPQ) material and social deprivation indices. These indices were initially developed in 1990 to measure deprivation among Quebecers and Canadians within a small geographical area and have been used as a substitute for individual-level socioeconomic data when analysing and identifying health and health system inequities ^28^. These indices use socioeconomic indicators collected in the Canadian census, spanning factors including levels of educational status, employment status, mean income, and the proportion of people living alone, that are separated, divorced, or widowed, or in single-parent families. Each census distribution area is then assigned a score based on a composite measure of these indicators, which is then translated into quintiles, where the fifth quintile represents the most deprived distribution areas ^28^. Deprivation quintiles were linked to cohort members based on their census distribution area at study entry, as recorded in the CLR.

##### Rurality

A two-level urban/rural variable was derived using Local Health Areas meeting the Statistics Canada definition for rural or remote areas ^34^. This was determined based on cohort members’ residence at the start of the study period.

### Statistical Analysis

A descriptive summary table presenting the distribution of exposure variables and covariates among cases and controls is provided for both the 30-and 90-day exposure windows in Table 3 and Table 4.

**Table 3.** Baseline Characteristics of Cases and Controls in the 30-day Exposure Window Analysis of Mental Health Contacts and Recurrent Overdose, Among People with a History of a Depression Diagnosis, British Columbia, 2015-2024

|  | Total<br>*n* = 5,505 | Cases<br>*n* = 944 | Controls<br>*n* = 4,561 |
| --- | --- | --- | --- |
| **NUMBER OF MENTAL HEALTH-RELATED PC CONTACTS** |  |  |  |
| Mean (SD) | 0.28 (0.89) | 0.14 (0.52) | 0.3 (0.95) |
| Median (IQR) | 0 (0 - 0) | 0 (0 - 0) | 0 (0 - 0) |
| **ANY MENTAL HEALTH-RELATED PC CONTACT** |  |  |  |
| No | 4,610 (83.74) | 850 (90.04) | 3,760 (82.44) |
| Yes | 895 (16.26) | 94 (9.96) | 801 (17.56) |
| **SEX** |  |  |  |
| Male | 2,949 (53.57) | 577 (61.12) | 2,372 (52.01) |
| Female | 2,556 (46.43) | 367 (38.88) | 2,189 (47.99) |
| **AGE GROUP** |  |  |  |
| 19-29 | 1,680 (30.52) | 375 (39.72) | 1,305 (28.61) |
| 30-39 | 1,308 (23.76) | 243 (25.74) | 1,065 (23.35) |
| 40-49 | 1,061 (19.27) | 189 (20.02) | 872 (19.12) |
| 50-59 | 874 (15.88) | 106 (11.23) | 768 (16.84) |
| 60+ | 582 (10.57) | 31 (3.28) | 551 (12.08) |
| **HEALTH AUTHORITY** |  |  |  |
| Fraser | 1,072 (19.47) | 147 (15.57) | 925 (20.28) |
| Interior | 591 (10.74) | 74 (7.84) | 517 (11.34) |
| Northern | 918 (16.68) | 212 (22.46) | 706 (15.48) |
| Vancouver Coastal | 1,406 (25.54) | 239 (25.32) | 1,167 (25.59) |
| Vancouver Island | 1,518 (27.57) | 272 (28.81) | 1,246 (27.32) |
| **MATERIAL DEPRIVATION (QUINTILE)** |  |  |  |
| Q1 (least deprived) | 862 (15.66) | 129 (13.67) | 733 (16.07) |
| Q2 | 959 (17.42) | 150 (15.89) | 809 (17.74) |
| Q3 | 1,064 (19.33) | 192 (20.34) | 872 (19.12) |
| Q4 | 1,240 (22.52) | 206 (21.82) | 1,034 (22.67) |
| Q5 (most deprived) | 1,380 (25.07) | 267 (28.28) | 1,113 (24.40) |
| **SOCIAL DEPRIVATION (QUINTILE)** |  |  |  |
| Q1 (least deprived) | 657 (11.93) | 101 (10.70) | 556 (12.19) |
| Q2 | 782 (14.21) | 126 (13.35) | 656 (14.38) |
| Q3 | 924 (16.78) | 139 (14.72) | 785 (17.21) |
| Q4 | 1,251 (22.72) | 210 (22.25) | 1,041 (22.82) |
| Q5 (most deprived) | 1,891 (34.35) | 368 (38.98) | 1,523 (33.39) |
| **NO FIXED ADDRESS** |  |  |  |
| No | 5,292 (96.13) | 903 (95.66) | 4,389 (96.23) |
| Yes | 213 (3.87) | 41 (4.34) | 172 (3.77) |

*PC = primary care. SD = standard deviation. IQR = interquartile range.*
*Cases = individuals with a recurrent non-fatal overdose (NFOD) or fatal overdose (FOD) between 30-days and one year following their index overdose, ascertained via BC Coroners Service and BC Emergency Health Services data. Controls = individuals without a recurrent overdose between 30-days and 1-year following their index overdose.*

**Table 4.** Baseline Characteristics of Cases and Controls in the 90-day Exposure Window Analysis of Mental Health Contacts and Recurrent Overdose Rates, British Columbia, 2015-2024

|  | Total<br>*n* = 5,105 | Cases<br>*n* = 641 | Controls<br>*n* = 4,464 |
| --- | --- | --- | --- |
| **NUMBER OF MENTAL HEALTH-RELATED PC CONTACTS** |  |  |  |
| Mean (SD) | 0.61 (1.5) | 0.46 (1.09) | 0.64 (1.55) |
| Median (IQR) | 0 (0 - 1) | 0 (0 - 0) | 0 (0 - 1) |
| **ANY MENTAL HEALTH-RELATED PC CONTACT** |  |  |  |
| No | 3,669 (71.87) | 494 (77.07) | 3,175 (71.12) |
| Yes | 1,436 (28.13) | 147 (22.93) | 1,289 (28.88) |
| **SEX** |  |  |  |
| Male | 2,709 (53.07) | 389 (60.69) | 2,320 (51.97) |
| Female | 2,396 (46.93) | 252 (39.31) | 2,144 (48.03) |
| **AGE GROUP** |  |  |  |
| 19-29 | 1,531 (29.99) | 245 (38.22) | 1,286 (28.81) |
| 30-39 | 1,223 (23.96) | 173 (26.99) | 1,050 (23.52) |
| 40-49 | 978 (19.16) | 123 (19.19) | 855 (19.15) |
| 50-59 | 827 (16.20) | 77 (12.01) | 750 (16.80) |
| 60+ | 546 (10.70) | 23 (3.59) | 523 (11.72) |
| **HEALTH AUTHORITY** |  |  |  |
| Fraser | 1,002 (19.63) | 98 (15.29) | 904 (20.25) |
| Interior | 554 (10.85) | 51 (7.96) | 503 (11.27) |
| Northern | 813 (15.93) | 122 (19.03) | 691 (15.48) |
| Vancouver Coastal | 1,323 (25.92) | 172 (26.83) | 1,151 (25.78) |
| Vancouver Island | 1,413 (27.68) | 198 (30.89) | 1,215 (27.22) |
| **MATERIAL DEPRIVATION (QUINTILE)** |  |  |  |
| Q1 (least deprived) | 813 (15.93) | 93 (14.51) | 720 (16.13) |
| Q2 | 897 (17.57) | 108 (16.85) | 789 (17.67) |
| Q3 | 991 (19.41) | 135 (21.06) | 856 (19.18) |
| Q4 | 1,147 (22.47) | 139 (21.68) | 1,008 (22.58) |
| Q5 (most deprived) | 1,257 (24.62) | 166 (25.90) | 1,091 (24.44) |
| **SOCIAL DEPRIVATION (QUINTILE)** |  |  |  |
| Q1 (least deprived) | 609 (11.93) | 65 (10.14) | 544 (12.19) |
| Q2 | 724 (14.18) | 84 (13.10) | 640 (14.34) |
| Q3 | 866 (16.96) | 96 (14.98) | 770 (17.25) |
| Q4 | 1,164 (22.80) | 141 (22.00) | 1,023 (22.92) |
| Q5 (most deprived) | 1,742 (34.12) | 255 (39.78) | 1,487 (33.31) |
| **NO FIXED ADDRESS** |  |  |  |
| No | 4,910 (96.18) | 611 (95.32) | 4,299 (96.30) |
| Yes | 195 (3.82) | 30 (4.68) | 165 (3.70) |

PC = primary care. SD = standard deviation. IQR = interquartile range.
Cases = individuals with a recurrent non-fatal overdose (NFOD) or fatal overdose (FOD) between 90-days and one year following their index overdose, ascertained via BC Coroners Service and BC Emergency Health Services data. Controls = individuals without a recurrent overdose between 30-days and 1-year following their index overdose.

**Table 3.**
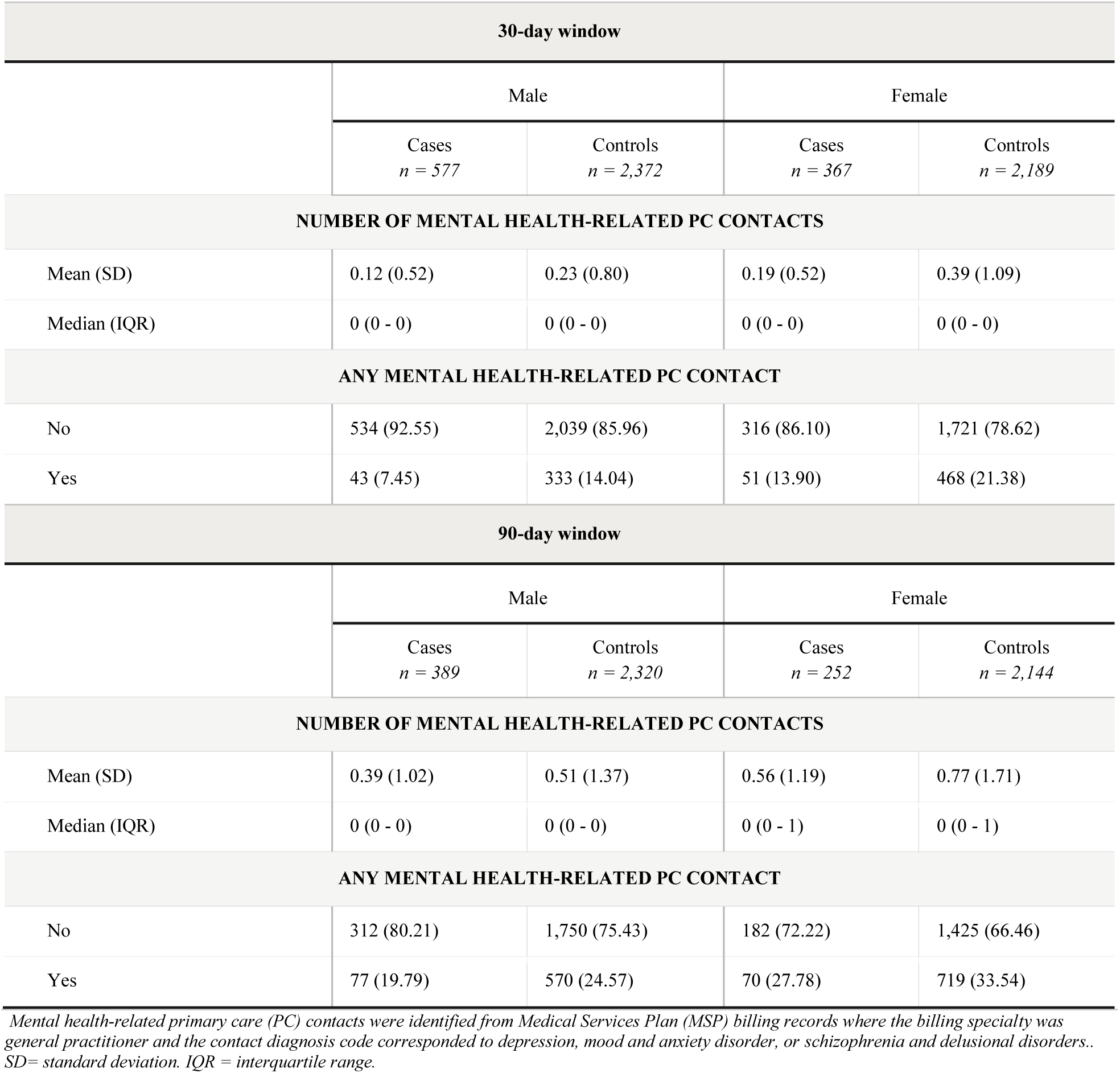
Distribution of Primary Care Contacts for Mental Health within 30- and 90-day Exposure Windows Among People with a History of a Depression Diagnosis, by Case-Control Status and Sex, British Columbia, 2015-2024

**Table 4.**
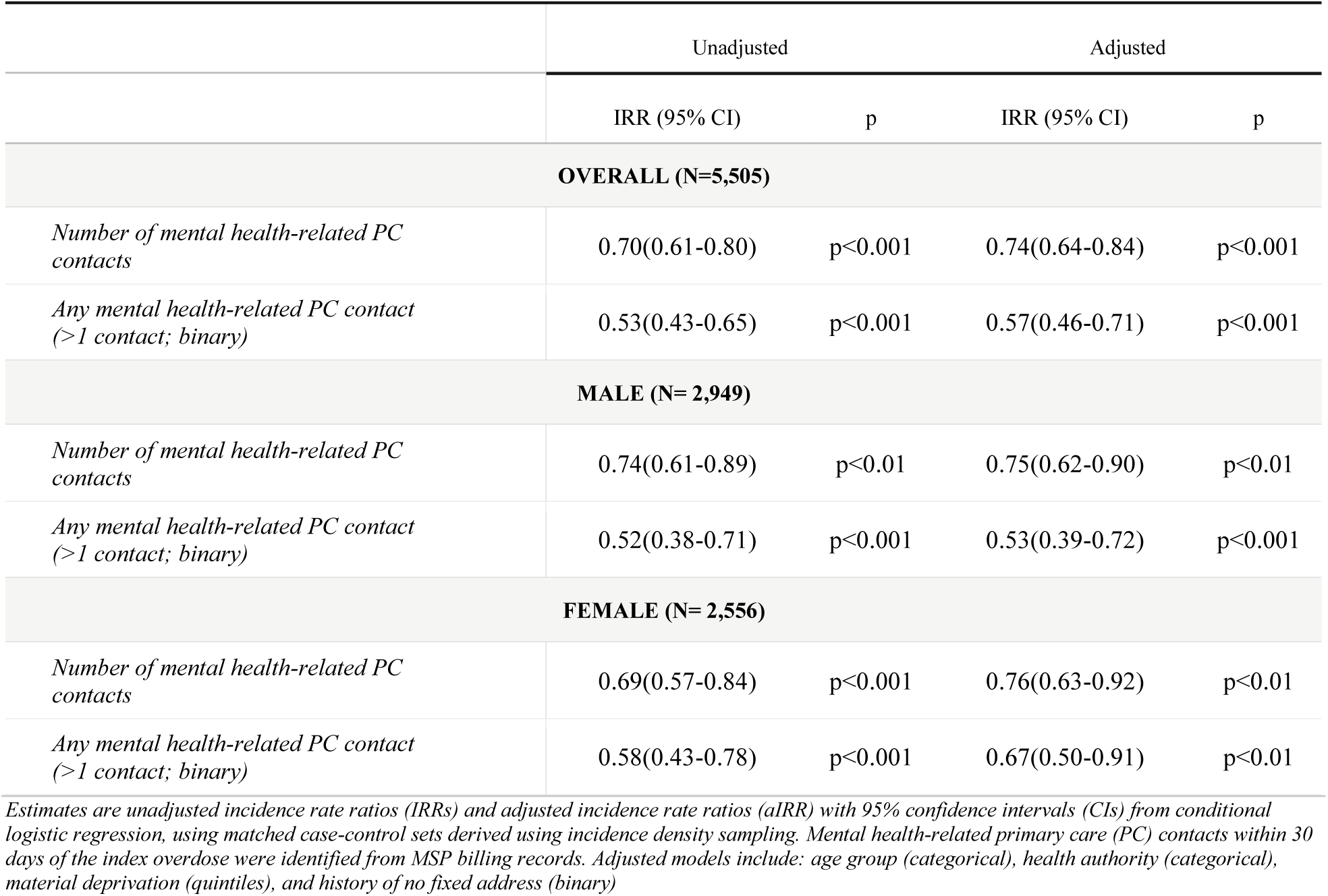
Association Between Mental Health-Related Primary Care Contacts Within 30-days Post Overdose and Rates of Recurrent Overdose Among People with a History of a Depression Diagnosis, Overall and Stratified by Sex, British Columbia, 2015-2024

Conditional logistic regression models were used to estimate the association of each mental health related primary care contact exposure variable with recurrent overdose risk, overall and stratified by sex. Models were first estimated without any adjustment variables (unadjusted), then with adjustment for age group, health authority, material deprivation, and no fixed address (adjusted). Models were also estimated using a rural/urban variable in place of the health authority variable, as well as an interaction term for material deprivation and housing status. Neither the rural/urban variable nor the interaction term for material deprivation and housing status improved model fit or altered exposure estimates, therefore, these results were not presented in the main analysis but are presented in Appendix A1. A model was also tested including the Elixhauser comorbidity index, excluding depression, but it did not meaningfully alter exposure estimates. Consistent with the incidence density sampling design, odds ratios ensuing from the conditional logistic regression models are interpreted as incidence rate ratios (IRRs)^35^.

To assess whether study findings were specific to primary care or reflected a broader pattern of mental health service contacts, a secondary analysis was conducted replacing the primary care exposure with contacts with a psychiatrist. These contacts were ascertained using the same procedure outlined in Section 3.2.4.1, using physician billing codes indicating the specialty of psychiatry.

### Sensitivity Analyses

Three sensitivity analyses were conducted. The first extended the follow-up period from the index overdose to two years. The second extended the follow-up period from the index overdose to the end of the study period (Dec 31, 2024). Finally, models were estimated using an unconditional logistic regression with adjustment for the matching variable, i.e. risk time, using a natural cubic spline ^36,37^.

## Results

### Sample Characteristics

Table 3 presents characteristics of cases and controls used for analyses of 30-day exposure periods. Overall, a low proportion of the entire study cohort had a mental health contact within 30 (16.26%) and 90 (28.13%) days. A higher proportion of controls (17.56%) than cases (9.96%) had at least one mental health related primary care contact within the 30-day period, and the mean number of contacts was higher among controls (mean: 0.30, SD: 0.95) than among cases (mean: 0.14, SD: 0.52). A similar pattern was observed for the 90-day exposure window (Table 4), with a higher proportion of controls (28.88%) than cases (22.93%) having any contact, and a higher mean number of contacts among controls (mean: 0.64, SD: 1.55) than cases (0.46, SD: 1.09). For both exposure windows, the distribution of the number of contacts was right-skewed (see Figures 3.4 and 3.5). For both 30- and 90-day windows, males comprised most of the study sample, and the proportion of females was lower among cases than controls.

Cases were more likely than controls to be in the highest deprivation quintiles. In the 30-day window 28.28% (material) and 38.98% (social) of cases were in the fifth quintile, representing the highest level of deprivation, compared to 24.40% and 33.39% among controls; a similar pattern was observed in the 90-day window. Cases were also more likely to reside in the Northern Health Authority (30-day: 22.46% of cases, 15.48% of controls; 90-day: 19.03% of cases, 15.48% of controls) and to be in the 19-29 age group (30-day cases: 39.72%, controls: 28.61%; 90-day cases: 38.22%, controls: 28.81%) and less likely to be in the 60+ age group (30-day cases: 3.28%, controls: 12.08%; 90-day cases: 3.59%, controls: 11.72%).

Table 5 presents the distribution of the exposure variables by sex. Females had higher rates of primary care contacts than males across both exposure windows and among both cases and controls. Among females in both exposure windows, cases were less likely than controls to have healthcare contacts (30-day window: 13.90% of cases versus 21.38% of controls; 90-day window: 27.78% of cases versus 33.54% of controls). Among males there was a similar pattern observed of cases having a lower proportion of contacts than controls (30-day window: 7.45% versus 14.04%; 90-day window: 19.79% and 24.57%), though the overall proportion of cases and controls with any contact was lower than among females.

**Table 5.** Association Between Mental Health-Related Primary Care Contacts within 90-days Post Overdose and Rates of Recurrent Overdose Among People with a History of a Depression Diagnosis, Overall and Stratified by Sex, British Columbia, 2015-2024

|  | Unadjusted |  | Adjusted<sup>a</sup> |  |
| --- | --- | --- | --- | --- |
|  | IRR (95% CI) | p | IRR (95% CI) | p |
| **OVERALL (N=5,105)** |  |  |  |  |
| *Number of mental health-related PC contacts* | 0.90(0.84-0.97) | p<0.01 | 0.92(0.86-0.99) | p<0.05 |
| *Any mental health-related PC contact (>1 contact; binary)* | 0.73(0.61-0.88) | p<0.01 | 0.78(0.65-0.94) | p<0.01 |
| **MALE (N=2,709)** |  |  |  |  |
| *Number of mental health-related PC contacts* | 0.92(0.83-1.01) | n.s. | 0.92(0.83-1.02) | n.s. |
| *Any mental health-related PC contact (>1 contact; binary)* | 0.76(0.60-0.98) | p<0.05 | 0.77(0.60-0.99) | p<0.05 |
| **FEMALE (N=2,396)** |  |  |  |  |
| *Number of mental health-related PC contacts* | 0.90(0.82-1.00) | n.s. | 0.94(0.86-1.04) | n.s. |
| *Any mental health-related PC contact (>1 contact; binary)* | 0.75(0.57-0.99) | p<0.05 | 0.84(0.63-1.11) | n.s. |

Estimates are unadjusted incidence rate ratios (IRRs) and adjusted incidence rate ratios (aIRR) with 95% confidence intervals (CIs) from conditional logistic regression, using matched case-control sets derived using incidence density sampling. Mental health-related primary care (PC) contacts within 90 days of the index overdose were identified from MSP billing records. Adjusted models include: age group (categorical), health authority (categorical), material deprivation (quintiles), and history of no fixed address (binary).

### Unadjusted Associations between Primary Care Contacts and Recurrent Overdose

#### Overall Analyses

Table 6 and Table 7 present unadjusted incidence rate ratios (IRR) and confidence intervals (CIs) for the association between mental health related primary care contact and rates of recurrent overdose, overall and stratified by sex (full model coefficients can be found in Appendix A.2 and Appendix A.3). Overall, mental health related primary care contacts were associated with lower rates of recurrent overdose with stronger associations observed for the 30-day exposure window than for the 90-day exposure window; having any mental health related primary care contact within 30 days was associated with 0.53 (CI: 0.43 - 0.65) times the rate of recurrent overdose, compared to 0.73 (CI: 0.61 - 0.88) times the rate for any contact within 90 days. Similarly, each mental health-related primary care contact within 30 days was associated with 0.70 (CI: 0.61 - 0.81) times the rate of recurrent overdose, compared to 0.90 (CI: 0.84 - 0.97) times the rate for each contact within 90 days.

**Table 6.** Secondary Analysis: Association Between Mental Health-Related Psychiatrist Contacts within 30-days Post Overdose and Rates of Recurrent Overdose Among People with a History of a Depression Diagnosis, Overall and Stratified by Sex, British Columbia, 2015-2024

|  | Unadjusted |  | Adjusted<sup>a</sup> |  |
| --- | --- | --- | --- | --- |
|  | IRR (95% CI) | p | IRR (95% CI) | p |
| **OVERALL (N=5,505)** |  |  |  |  |
| *Number of mental health-related Psychiatrist contacts* | 0.87(0.82-0.92) | p<0.001 | 0.89(0.85-0.94) | p<0.001 |
| *Any mental health-related Psychiatrist contact (>1 contact; binary)* | 0.51(0.40-0.65) | p<0.001 | 0.59(0.47-0.75) | p<0.001 |
| **MALE (N= 2,949)** |  |  |  |  |
| *Number of mental health-related Psychiatrist contacts* | 0.93(0.88-0.98) | p<0.05 | 0.94(0.89-1.00) | p<0.05 |
| *Any mental health-related Psychiatrist contact (>1 contact; binary)* | 0.65(0.48-0.87) | p<0.01 | 0.70(0.52-0.95) | p<0.05 |
| **FEMALE (N= 2,556)** |  |  |  |  |
| *Number of mental health-related Psychiatrist contacts* | 0.77(0.68-0.88) | p<0.001 | 0.81(0.72-0.91) | p<0.001 |
| *Any mental health-related Psychiatrist contact (>1 contact; binary)* | 0.41(0.28-0.59) | p<0.001 | 0.50(0.34-0.73) | p<0.001 |

Estimates are unadjusted incidence rate ratios (IRRs) and adjusted incidence rate ratios (aIRR) with 95% confidence intervals (CIs) from conditional logistic regression, using matched case-control sets derived using incidence density sampling. Mental health-related psychiatrist contacts within 30 days of the index overdose were identified from MSP billing records. Adjusted models include: age group (categorical), health authority (categorical), material deprivation (quintiles), and history of no fixed address (binary)

**Table 7.** : Secondary Analysis: Association Between Mental Health-Related Psychiatrist Contacts within 90-days Post Overdose and Rates of Recurrent Overdose Among People with a History of a Depression Diagnosis, Overall and Stratified by Sex, British Columbia, 2015-2024

|  | Unadjusted |  | Adjusted<sup>a</sup> |  |
| --- | --- | --- | --- | --- |
|  | IRR (95% CI) | p | IRR (95% CI) | p |
| **OVERALL (N=5,105)** |  |  |  |  |
| *Number of mental health-related Psychiatrist contacts* | 0.96(0.93-0.99) | p<0.01 | 0.97(0.94-1.00) | p<0.05 |
| *Any mental health-related Psychiatrist contact (>1 contact; binary)* | 0.61(0.48-0.77) | p<0.001 | 0.67(0.53-0.85) | p<0.01 |
| **MALE (N=2,709)** |  |  |  |  |
| *Number of mental health-related Psychiatrist contacts* | 0.99(0.96-1.01) | n.s. | 0.99(0.96-1.02) | n.s. |
| *Any mental health-related Psychiatrist contact (>1 contact; binary)* | 0.75(0.55-1.01) | n.s. | 0.77(0.57-1.04) | n.s. |
| **FEMALE (N=2,396)** |  |  |  |  |
| *Number of mental health-related Psychiatrist contacts* | 0.91(0.85-0.97) | p<0.01 | 0.92(0.87-0.99) | p<0.05 |
| *Any mental health-related Psychiatrist contact (>1 contact; binary)* | 0.50(0.34-0.73) | p<0.001 | 0.60(0.41-0.88) | p<0.01 |

Estimates are unadjusted incidence rate ratios (IRRs) and adjusted incidence rate ratios (aIRR) with 95% confidence intervals (CIs) from conditional logistic regression, using matched case-control sets derived using incidence density sampling. Mental health-related psychiatrist contacts within 90 days of the index overdose were identified from MSP billing records. Adjusted models include: age group (categorical), health authority (categorical), material deprivation (quintiles), and history of no fixed address (binary)

#### Sex-stratified analyses

In sex-stratified analyses, the association between having any mental health related primary care contact within 30 days and recurrent overdose was modestly stronger for males (IRR: 0.52, CI: 0.38 - 0.71) than it was for females (IRR: 0.58, CI: 0.43 - 0.78); conversely, the association between the number of primary care contacts for mental health within 30 days and recurrent overdose was modestly stronger for females (IRR: 0.69, CI: 0.57 - 0.84) than it was for males (IRR: 0.74, CI: 0.61 - 0.89). The pattern of associations being weaker for 90-day windows persisted in sex-stratified analyses. Among females, any contact within 90 days was associated with 0.75(CI: 0.57 - 0.99) times the rate of recurrent overdose, while each contact within 90 days was associated with 0.84 (CI: 0.63 – 1.11) times the rate; corresponding associations for males were similar, with any contact within 90 days associated with 0.76 (CI: 0.60 - 0.98) times the rate of recurrent overdose and each contact associated with 0.92 (CI: 0.83 - 1.01) times the rate of recurrent overdose. Of note, the number of contacts within 90 days was not statistically significantly associated with recurrent overdose for either males or females.

### Adjusted Associations between Primary Care Contacts and Recurrent Overdose

#### Overall Analyses

In overall analyses, adjusting for covariates had minimal attenuating effects on associations between mental health related primary care contacts and the rate of recurrent overdose (Table 6 and Table 7; see also Figures 6 and 7). Having any contact within 30 days was associated with 0.57 (CI: 0.46 - 0.71) times the rate of recurrent overdose, and each contact within 30 days was associated with 0.74 (CI: 0.64 - 0.84) times the rate; corresponding IRRs for the 90-day window were 0.78 (CI: 0.65 - 0.94) for any contact and 0.92 (CI: 0.86 - 0.99) for the number of contacts.

**Figure 6.**
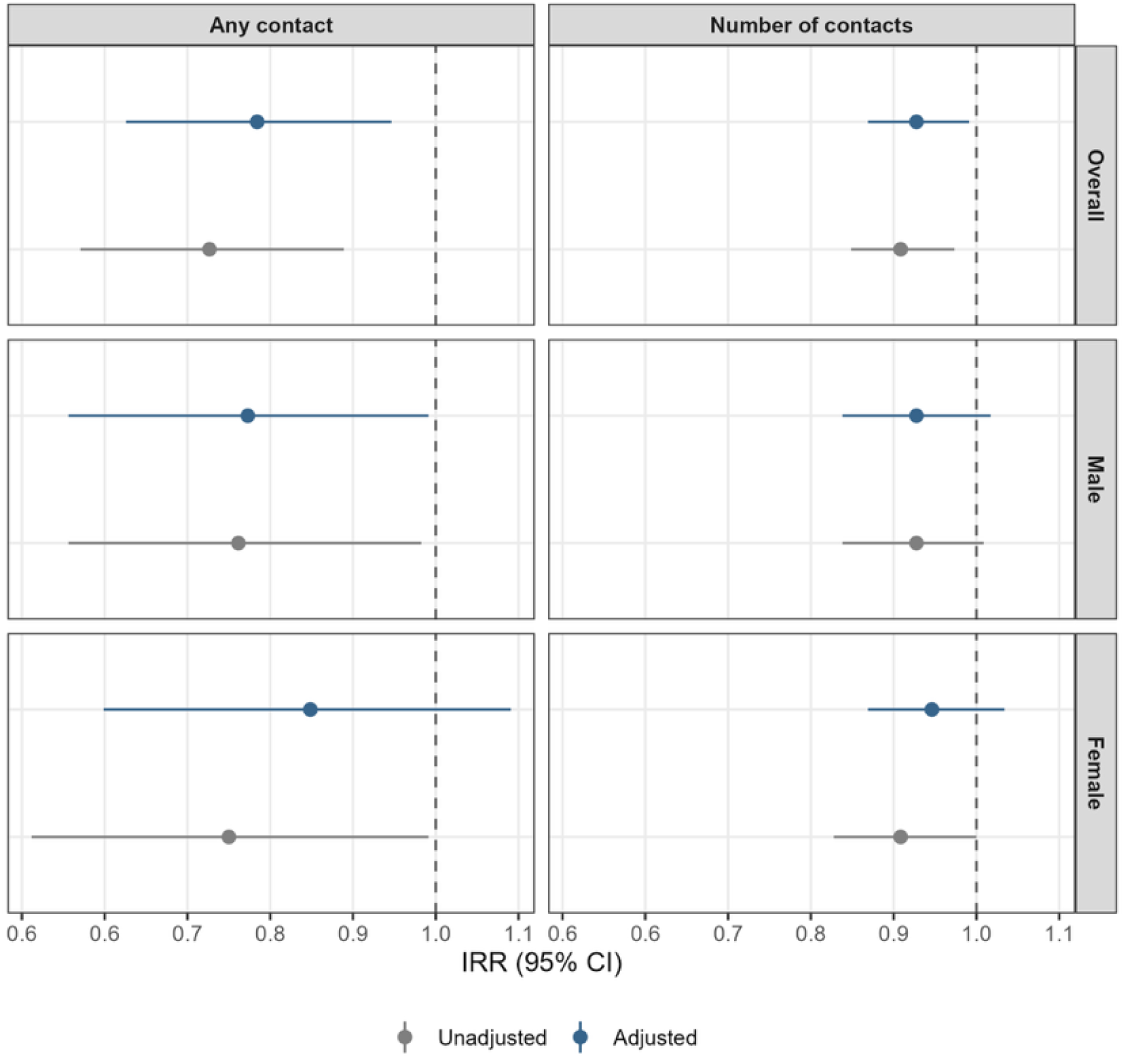
Forest Plot of Incidence Rate Ratios and 95% Confidence Intervals for Mental Health-Related Primary Care Contacts within 90-days Post Overdose and Rates of Recurrent Overdose Among People with a History of a Depression Diagnosis.

#### Sex-stratified analyses

In sex-stratified analyses, adjusting for covariates produced more notable attenuation of the associations between primary care contacts for mental health contacts and recurrent overdose rates among females than among males. Among females, having any contact within 30 days was associated with 0.67 (CI: 0.50 - 0.91) times the rate of recurrent overdose, and having any contact with 90 days was associated with 0.84 (CI: 0.63 - 1.11) the rate; each contact within 30 days was associated with 0.76 (CI: 0.63 - 0.92) times the rate of recurrent overdose, and each contact within 90 days was associated with 0.94 (CI: 0.86 - 1.04) times the rate of recurrent overdose. Among males the adjusted incidence rate ratios were largely consistent with the unadjusted incidence rate ratios; any contact within 30 or 90 days was associated with 0.53 (CI: 0.39 - 0.72) and 0.77 (CI: 0.60 - 0.99) times the rate of recurrent overdose, and each contact within 30 or 90 days was associated with 0.75 (CI: 0.62 - 0.90) and 0.92 (CI: 0.83 - 1.02) times the rate of recurrent overdose.

The greater attenuation among females, relative to the minimal attenuation observed among males, affected the sex differences observed in the unadjusted analyses differently depending on the exposure variable. For measures of any contact within 30 or 90 days, where the association between having any contact and the rate of recurrent overdose was already stronger among males than females in unadjusted estimates, adjustment further increased this difference. Meanwhile, where the association between the number of contacts and the rate of recurrent overdose was stronger among females than males in unadjusted estimates, adjustment brought the estimates among females closer to those for males, resulting in similar aIRRs for both sexes. Of note, in the sex stratified analyses estimated adjusted incidence rate ratios for the number of contacts within 90 days were not statistically significant for either sex, and the adjusted rate ratio for any primary care contact within 90 days was not statistically significant for females; this in contrast to the analyses within the 30-day exposure window, where all estimates were statistically significant.

### Secondary Analysis: Psychiatrist Contacts for Mental Health as the Exposure

#### Overall Analyses

Secondary analyses using psychiatrist contacts as the mental health exposure showed protective associations against recurrent overdose, consistent with the primary analyses in Sections 3.3.2 and 3.3.3. Table 8 and Table 9 presents IRRs and aIRRs for the association between psychiatrist contacts and recurrent overdose, overall and stratified by sex. In overall analyses, having any psychiatrist contact within 30 days was associated with 0.51 (CI: 0.40 - 0.65) times the rate of recurrent overdose (aIRR: 0.59, CI: 0.47 - 0.75), and any contact within 90 days was associated with 0.61 (CI: 0.48 - 0.77) times the rate of recurrent overdose (aIRR: 0.67, CI: 0.53 - 0.85); these associations were stronger than the corresponding primary care estimates. Conversely, associations for the number of psychiatrist contacts were weaker than for the number of primary care contacts for both the 30-day (IRR: 0.87, CI: 0.82 - 0.92; aIRR: 0.89, CI: 0.85 - 0.94) and 90-day (IRR: 0.96, CI: 0.93 - 0.99; aIRR: 0.97, CI: 0.94 - 1.00) windows. Adjusting for covariates had a greater attenuating effect on the associations between any psychiatrist contact within 30 or 90 days and recurrent overdose rates compared to the corresponding estimates for primary care contacts.

**Table 8.** Distribution of Mental Health-Related Psychiatrist Contacts within 30- and 90-Day Exposure Windows Among People with a History of a Depression Diagnosis by Sex, among Cases and Controls, British <u>Columbia, 2015-2024</u>

| 30-day window |  |  |  |  |
| --- | --- | --- | --- | --- |
|  | Male |  | Female |  |
|  | Cases<br>*n* = 577 | Controls<br>*n* = 2,372 | Cases<br>*n* = 367 | Controls<br>*n* = 2,189 |
| NUMBER OF MENTAL HEALTH-RELATED PSYCHIATRIST CONTACTS |  |  |  |  |
| Mean (SD) | 0.28 (1.58) | 0.56 (2.34) | 0.18 (0.93) | 0.85 (2.78) |
| Median (IQR) | 0 (0 – 0) | 0 (0 – 0) | 0 (0 – 0) | 0 (0 – 0) |
| ANY MENTAL HEALTH-RELATED PSYCHIATRIST CONTACT |  |  |  |  |
| No | 531 (92.03) | 2,076 (87.52) | 338 (92.10) | 1,795 (82.00) |
| Yes | 46 (7.97) | 296 (12.48) | 29 (7.90) | 403 (18.00) |
| 90-day window |  |  |  |  |
|  | Male |  | Female |  |
|  | Cases<br>*n* = 389 | Controls<br>*n* = 2,320 | Cases<br>*n* = 252 | Controls<br>*n* = 2,144 |
| NUMBER OF MENTAL HEALTH-RELATED PSYCHIATRIST CONTACT |  |  |  |  |
| Mean (SD) | 0.83 (3.51) | 1.01 (4.18) | 0.48 (2.01) | 1.47 (5.20) |
| Median (IQR) | 0 (0 – 0) | 0 (0 – 0) | 0 (0 – 0) | 0 (0 – 0) |
| ANY MENTAL HEALTH-RELATED PSYCHIATRIST CONTACT |  |  |  |  |
| No | 340 (87.40) | 1,938 (83.53) | 221 (87.70) | 1,663 (77.57) |
| Yes | 49 (12.60) | 382 (16.47) | 31 (12.30) | 481 (22.43) |

**Table 9.**
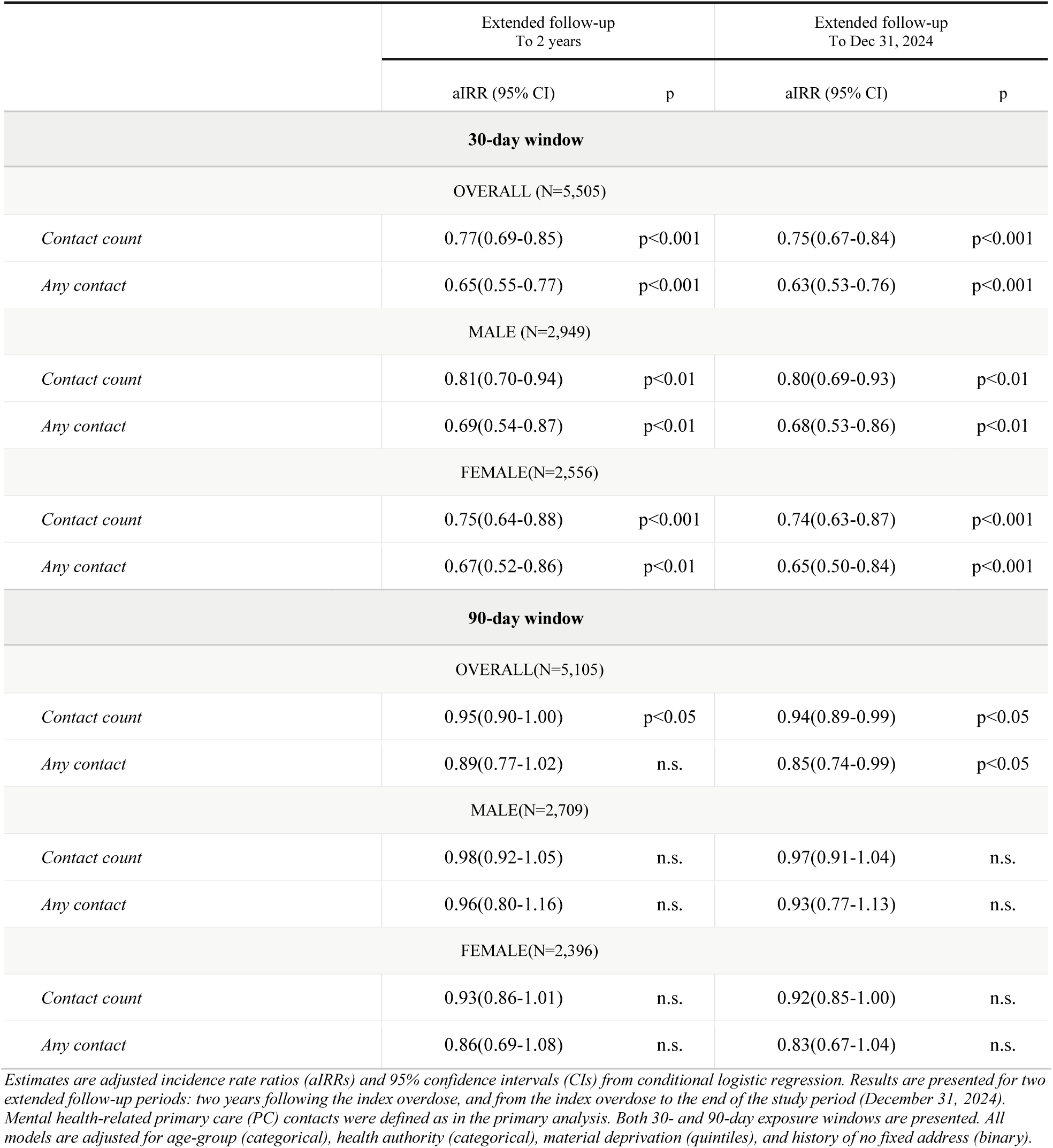
Sensitivity Analysis: Association Between Mental Health-Related Primary Care Contacts and Rates of Recurrent Overdose Among People with a History of a Depression diagnosis Under Extended Follow-Up Periods, British Columbia, 2015-2024

|  | Extended follow-up<br>To 2 years |  | Extended follow-up<br>To Dec 31, 2024 |  |
| --- | --- | --- | --- | --- |
|  | aIRR (95% CI) | p | aIRR (95% CI) | p |
| **30-day window** |  |  |  |  |
| OVERALL (N=5,505) |  |  |  |  |
| *Contact count* | 0.77(0.69-0.85) | p<0.001 | 0.75(0.67-0.84) | p<0.001 |
| *Any contact* | 0.65(0.55-0.77) | p<0.001 | 0.63(0.53-0.76) | p<0.001 |
| MALE (N=2,949) |  |  |  |  |
| *Contact count* | 0.81(0.70-0.94) | p<0.01 | 0.80(0.69-0.93) | p<0.01 |
| *Any contact* | 0.69(0.54-0.87) | p<0.01 | 0.68(0.53-0.86) | p<0.01 |
| FEMALE(N=2,556) |  |  |  |  |
| *Contact count* | 0.75(0.64-0.88) | p<0.001 | 0.74(0.63-0.87) | p<0.001 |
| *Any contact* | 0.67(0.52-0.86) | p<0.01 | 0.65(0.50-0.84) | p<0.001 |
| **90-day window** |  |  |  |  |
| OVERALL(N=5,105) |  |  |  |  |
| *Contact count* | 0.95(0.90-1.00) | p<0.05 | 0.94(0.89-0.99) | p<0.05 |
| *Any contact* | 0.89(0.77-1.02) | n.s. | 0.85(0.74-0.99) | p<0.05 |
| MALE(N=2,709) |  |  |  |  |
| *Contact count* | 0.98(0.92-1.05) | n.s. | 0.97(0.91-1.04) | n.s. |
| *Any contact* | 0.96(0.80-1.16) | n.s. | 0.93(0.77-1.13) | n.s. |
| FEMALE(N=2,396) |  |  |  |  |
| *Contact count* | 0.93(0.86-1.01) | n.s. | 0.92(0.85-1.00) | n.s. |
| *Any contact* | 0.86(0.69-1.08) | n.s. | 0.83(0.67-1.04) | n.s. |

Estimates are adjusted incidence rate ratios (aIRRs) and 95% confidence intervals (CIs) from conditional logistic regression. Results are presented for two extended follow-up periods: two years following the index overdose, and from the index overdose to the end of the study period (December 31, 2024). Mental health-related primary care (PC) contacts were defined as in the primary analysis. Both 30- and 90-day exposure windows are presented. All models are adjusted for age-group (categorical), health authority (categorical), material deprivation (quintiles), and history of no fixed address (binary).

#### Sex-stratified Analyses

In sex-stratified analyses, and contrary to the findings for primary care contacts, associations between the contact variables and recurrent overdose were stronger for females than for males (Table 8 and Table 9). Specifically, any psychiatrist contact within 30 days was associated with 0.41 (CI: 0.28 - 0.59) the rate of recurrent overdose among females compared to 0.65 (CI: 0.48 - 0.87) among males, while any psychiatrist contact within 90 days was associated with 0.50 (CI: 0.34 - 0.73) the rate among females compared to 0.75 (CI: 0.55 - 1.01) among males. Corresponding IRRs for the number of psychiatrist contacts were 0.77 (CI: 0.68 - 0.88) and 0.91 (CI: 0.85 - 0.97) for 30 and 90 days among females, compared to 0.93 (CI: 0.88 - 0.98) and 0.99 (CI: 0.96 - 1.01) among males.

After adjustment, associations remained stronger among females than among males; aIRRs for any psychiatrist contact within 30 or 90 days were 0.50 (CI: 0.34 - 0.73) and 0.60 (CI: 0.41 - 0.88) among females compared to 0.70 (CI: 0.52 - 0.95) and 0.77 (CI: 0.57 - 1.04) among males. After adjustment, the associations between the number of psychiatrist contacts and recurrent overdose risk continued to be weaker than for primary care contacts, with IRRs for the number of contacts within 30 and 90 days were 0.81 (CI: 0.72 - 0.91) and 0.92 (CI: 0.87 - 0.99) among females and 0.94 (CI: 0.89 - 1.00) and 0.99 (CI: 0.96 - 1.02) among males. Notably, estimated aIRRs for any psychiatrist contact and the number of psychiatrist contacts within 90 days were both not statistically significant among males.

Of note, the underlying distribution of psychiatrist contacts differed from that of primary care contacts, as seen in Table 10. Specifically, sex differences in the distribution of psychiatrist contacts differed from those seen in primary care: female cases had a similar proportion with any psychiatrist contact for both 30 (7.90%) and 90 (12.30%) day windows to males (30 day: 7.97%; 90 day: 12.60%), while female controls had higher proportions with any contact in both windows (30 days: 18.00%, 90 days: 22.43%) compared to males (30 days: 12.48%, 90 days: 16.47%). Despite similar proportions, male cases had a higher mean number of psychiatrist contacts than female cases for both 30 (male: 0.28, SD: 1.58; female: 0.18, SD: 0.93) and 90-day windows (male: 0.83, SD: 3.51; female: 0.48, SD: 2.01).

### Sensitivity Analyses

#### Extended Follow-Up Time for Ascertainment of Recurrent Overdose

Sensitivity analyses extending the follow-up time showed protective associations for mental-health related primary care contacts within 30 days of overdose, consistent with findings in Section 3.3.3, Table 11 presents the adjusted incidence rate ratios for the association between mental health related primary care contact and recurrent overdose for analyses within extended follow-up periods. Extending the follow-up period attenuated the estimated aIRRs. In the overall analyses, extending the follow-up to two years attenuated the aIRR for any contact within 30 days to 0.65 (CI: 0.55 -0.77) and for number of contacts within 30 days to 0.77 (CI: 0.69 - 0.85), with the corresponding estimates for the 90-day window of 0.89 (CI: 0.77 - 1.02) and 0.95 (CI: 0.90 - 1.00). When extending the follow-up time to the end of the study period (December 31, 2024), the aIRRs for any mental health related primary care contact and the number of contacts within 30 days were 0.63 (CI: 0.53 - 0.76) and 0.75 (CI: 0.67 - 0.84), while for 90 days corresponding estimates were 0.85 (CI: 0.74 - 0.99) and 0.94 (CI: 0.89 - 0.99). In contrast to the main analyses, sex-specific adjusted rate ratios were notably similar in both extended follow-up analyses. For instance, for the estimates of the association between any contact within 30 days, where sex differences were pronounced in the main analyses, sex specific estimates in the analyses using two year follow-up for males (aIRR: 0.69, CI: 0.54 - 0.87) and females (aIRR: 0.67, CI: 0.52 - 0.86) were similar, which was also reflected for the estimates for males (aIRR: 0.68, CI: 0.53 - 0.86) and females (aIRR: 0.65, CI: 0.50 - 0.84) for analyses using the full study period follow-up.

#### Analyses Using Unconditional Logistic Regression Models

The unconditional logistic regression sensitivity analysis produced results consistent with the estimates presented for the results in 3.3, with aIRR estimates in these models within 0.01 of the aIRR estimates presented in the main analysis. These results can be found in Appendix A4.

## Discussion

This study found that contacts with a primary care provider for mental health have a protective association with recurrent overdose risk among those with a documented depression diagnosis history; these associations were strong and present for both males and females, suggesting that mental health care, including in primary care settings, should be considered an important component of preventing longer term recurrence of overdose. Contacts with primary care for mental health in the 30-day window had stronger protective associations with overdose recurrence than in the 90-day window, suggesting that timeliness of care may be a key factor for reducing recurrent overdose risk. This finding is strengthened by the sensitivity analyses with extended follow-up time, where the association with recurrence risk for exposures using the 30-day exposure window have persistent, albeit weakened relative to primary analyses, protective association with overdose, while the association with recurrence risk for exposures using the 90-day exposure window was not present with extended follow-up.

Despite these findings, care accessibility appeared to be a striking issue, given the low proportion of people with a documented depression history that accessed primary care for mental health following a non-fatal overdose. Unmeasured factors such as stigma could be dissuading people from seeking care^38^, and there remains approximately 20% of people in BC without an attached primary care provider (PCP) ^39^. Additionally, literature suggests that those with more complex health needs, such as a history of mental health and/or substance use conditions may face increased difficulty in accessing primary care, especially when facing social and structural vulnerabilities ^40–43^. These barriers may be further compounded by structural factors such as the accessibility of transportation to and from appointments ^41,44^, which may be exacerbated by the social and material deprivation experienced by a substantial proportion of the study cohort. Additionally, it is worth noting that the low rates of care contacts observed in this study occurred within a population that has inherently engaged with the health system sufficiently to receive a documented depression diagnosis.

This study found modest sex differences in the associations between mental health contacts and recurrent overdose risk. These differences varied in terms of direction and magnitude depending on the type of practitioner and the exposure window, with the protective associations between primary care contacts on recurrent overdose risk being modestly stronger for males than females in the 30-day window, and to a lesser extent in the 90-day window, though the reverse was observed for the protective associations between psychiatrist contacts on recurrent overdose risk were stronger among females than males. Sex differences seemed to dissipate as follow-up time was extended, though this could be in part due to the weaker observed associations in extended follow-up time overall.

The reversal in the direction of sex differences between provider types may reflect differences in the post-overdose needs of males and females. Prior research on women and females in substance use treatment indicates higher rates of co-occurring mental health conditions, trauma histories, and other psychosocial vulnerabilities, which may require or benefit from more specialized care than is available in a primary care setting^13,14^. Additionally, the higher mean number of psychiatrist contacts observed among male cases relative to female cases, despite relatively similar proportions having any contact, may also reflect sex differences in the intensity and type of treatment received.

Overall, males seemed to access care less than females as is consistent with a significant body of research on men’s and males’ health and health-seeking behaviours^45,46^. This pattern likely reflects an interplay between individual factors, such as gendered norms surrounding help-seeking, and interpersonal factors, such as differences in types of treatment received for depression by males and females ^47^. An ongoing need, then, is improving males’ access to and engagement with care given the sustained disproportionate rates of both fatal and non-fatal overdose among males. This is particularly notable given the aforementioned inherent level of contact with care among people in this study, suggesting that care engagement gaps are not merely a product of health system entry.

The greater attenuation of the association between primary care contacts for mental health and overdose recurrence among females observed may indicate that social and structural vulnerabilities may more substantially influence overdose risk among females than males. Consistent with the socioecological model, the impact of interpersonal factors such as contacts with healthcare may be altered by structural conditions that disproportionally affect different segments of the population ^33^. Taken together, these findings indicate that while mental health care in both primary care and specialist settings have protective associations against recurrent overdose, mechanisms and moderating factors thereof may differ by sex. As such, while this study in isolation does not strongly indicate a need for sex differentiation in interventions for the prevention of recurrent overdose, it may suggest that further investigation into sex differences in post-overdose care more broadly are warranted. Specifically, potential sex differences in referral pathways and patterns of accessing care between the sexes could be an important area for further investigation of sex differences, especially in the context of research on other mental health and substance use interventions, such as antidepressant initiation ^12–14,47^, which have found sex differences in the likelihood of receiving these types of care and the effects thereof.

### Strengths

This study used a nested-care control design with incidence density sampling, which both accounts for person-time at risk and allows for the testing of multiple different exposure variables, providing the opportunity for a more detailed analysis of the association between primary care for mental health on overdose risk than study designs that only allow for the testing of one exposure variable. The use of multiple linked administrative health datasets provided relatively robust data on health system contacts, overdoses, and some sociodemographic characteristics across the entire population of interest, specifically people in BC with a documented history of a depression diagnosis who have experienced an overdose. As well, this study captured a substantial proportion of mental health care received by this population, insofar as it captured primary care, which is the most common site where people in Canada receive mental health care ^48^, while also capturing specialist care in the form of psychiatrist care. The inclusion of a secondary analysis examining psychiatrist contacts for mental health as an alternate exposure further strengthened the study by allowing for comparisons across different provider types and provided insight into whether the observed associations are specific to primary care or are reflective of mental health care more broadly. Finally, the use of multiple sensitivity analyses varying different components of the study design, including the type of healthcare provider, follow-up length, and regression specification, provided additional context and confidence in the main findings.

### Limitations

Several limitations of this study should be noted. Skew in exposure variables, specifically the number of contacts, could affect the reliability of the estimates, even with the high number of controls per case. The generalizability of study findings is somewhat limited by the study’s scope and data limitations. The study only captured a subset of people with a documented history of a depression diagnosis, who therefore inherently have accessed at least some medical care, and have had a non-fatal overdose for which they received care, meaning this study’s findings may not reflect those who have not engaged with the health system, and may not reflect the broader population of people who have experienced an overdose but do not have a documented depression diagnosis history. As well, this study did not capture mental health care received outside of the provincial healthcare system, such as private counselling.

The exclusion of cases within 30 and 90 days likely excluded those at highest risk of recurrent overdose, which could lead to overestimating the strength of the association between the care contact variables and recurrent overdose risk. While this study was focused less on longer-term follow-up care and risk of recurrent overdose, this nonetheless presents a substantial limitation that these people are not included in this analysis. Additionally, mental health service contacts included all mental health conditions, not just depression, to ensure that potentially relevant contacts were not missed due to variations in physician billing/coding. As a result, some of the contacts and ensuing associations may be due to the treatment of other underlying mental health conditions, such as severe mental illness, that could be associated with recurrent overdose risk.

The measurement of housing status in this study presents further limitations. No Fixed Address data only captures people with hospitalizations and emergency department visits, which limits the identification of those experiencing housing deprivation and precarity to people for whom this status was recorded by a health care provider and is available in administrative health records. This may also not capture people who are precariously housed, but do not disclose this or provide a friend or family member’s address as their own, and therefore do not get coded as having no fixed address. As well, people who are unhoused may be less likely to have their healthcare contacts and records linked in administrative data and therefore may be more likely to be missing in the data ^49,50^. Social assistance data can better identify those with no fixed address, but this data was not available at the time of analysis. With regards to deprivation more broadly, commonly used indices of deprivation in Canada, such as the INSPQ indices and the Canadian Index of Multiple Deprivation, may inadequately capture deprivation experienced by Indigenous people^51,52^. While this study did not have direct data on Indigenous identity, Indigenous people still exist in the data, and, even where their identity is not flagged as such, they continue to experience Indigenous-specific forms of deprivation. Therefore, the use of the INSPQ indices does not sufficiently adjust for this deprivation, since these indices rely on census indicators that do not capture social exclusion and differences in, for instance, education delivery on reserves that contribute to deprivation^51,52^.

Finally, covariates were measured at study entry (January 1, 2015) rather than the index or recurrent overdose. This approach was taken for both simplicity and to ensure consistency across cases and controls, given that people could be sampled at multiple times throughout the study period, given the incidence density sampling design. However, as some of these characteristics may change between study entry and the index or recurrent overdose, this approach may introduce misclassification bias for the covariate effects.

## Data Availability

Requests for access to the data in the present study can be made directly through correspondence with the corresponding author

## Acknowledgements

- Access to data provided by the Data Steward(s) is subject to approval, but can be requested for research projects through the Data Steward(s) or their designated service providers. All inferences, opinions, and conclusions drawn in this publication are those of the author(s), and do not reflect the opinion of the Data Steward(s). The following data sets were used in this project: Medical Services Plan (MSP), Discharge Abstract Database (DAD), BC Emergency Health Services (BCEHS), BC Coroners Service (BCCS), and Client Roster (CLR), National Ambulatory Care Reporting System (NACRS). This Data was provisioned under ISP 21-051.
- I gratefully acknowledge the residents of British Columbia whose data are integrated in the data sets we used on PANDA.
- I acknowledge the Professionals for the Ethical Engagement of Peers (PEEP) and First Nations Health Authority (FNHA) Health Surveillance team for their feedback on this work.

## Appendices

**A1.** Alternate Models Examining Association Between Mental Health-Related Primary Care Contacts and Rates of Recurrent Overdose Among People with a History of a Depression Diagnosis, British Columbia, 2015-2024.

|  | Models Using a Rural/Urban Term Instead of Health Authority |  | Models Using a No Fixed Address and Material Deprivation Interaction Term |  |
| --- | --- | --- | --- | --- |
|  | aIRR (95% CI) | p | aIRR (95% CI) | p |
| **30-day window** |  |  |  |  |
| OVERALL (N=5,505) |  |  |  |  |
| *Contact count* | 0.73(0.64-0.84) | p<0.001 | 0.74(0.64-0.84) | p<0.001 |
| *Any contact* | 0.57(0.46-0.70) | p<0.001 | 0.58(0.46-0.71) | p<0.001 |
| MALE (N=2,949) |  |  |  |  |
| *Contact count* | 0.74(0.61-0.90) | p<0.01 | 0.75(0.62-0.90) | p<0.01 |
| *Any contact* | 0.53(0.38-0.72) | p<0.001 | 0.53(0.39-0.72) | p<0.001 |
| FEMALE(N=2,556) |  |  |  |  |
| *Contact count* | 0.75(0.62-0.91) | p<0.01 | 0.77(0.64-0.92) | p<0.01 |
| *Any contact* | 0.66(0.49-0.89) | p<0.01 | 0.69(0.51-0.92) | p<0.05 |
| **90-day window** |  |  |  |  |
| OVERALL(N=5,105) |  |  |  |  |
| *Contact count* | 0.92(0.86-0.99) | p<0.05 | 0.92(0.86-0.99) | p<0.05 |
| *Any contact* | 0.77(0.64-0.93) | p<0.01 | 0.78(0.65-0.94) | p<0.01 |
| MALE(N=2,709) |  |  |  |  |
| *Contact count* | 0.92(0.83-1.02) | n.s. | 0.92(0.83-1.02) | n.s. |
| *Any contact* | 0.76(0.59-0.98) | p<0.05 | 0.77(0.60-1.00) | p<0.05 |
| FEMALE(N=2,396) |  |  |  |  |
| *Contact count* | 0.94(0.85-1.04) | n.s. | 0.94(0.86-1.04) | n.s. |
| *Any contact* | 0.83(0.63-1.09) | n.s. | 0.85(0.64-1.12) | n.s. |

aIRR = adjusted incidence rate ratio; CI= 95% confidence interval; n.s. = not statistically significant. Visist count refers to the total number of mental health related primary care contacts within the specified exposure window. Any contact refers to the presence of at least one mental health related primary care contact within the specified exposure window.

**Appendix A2.**
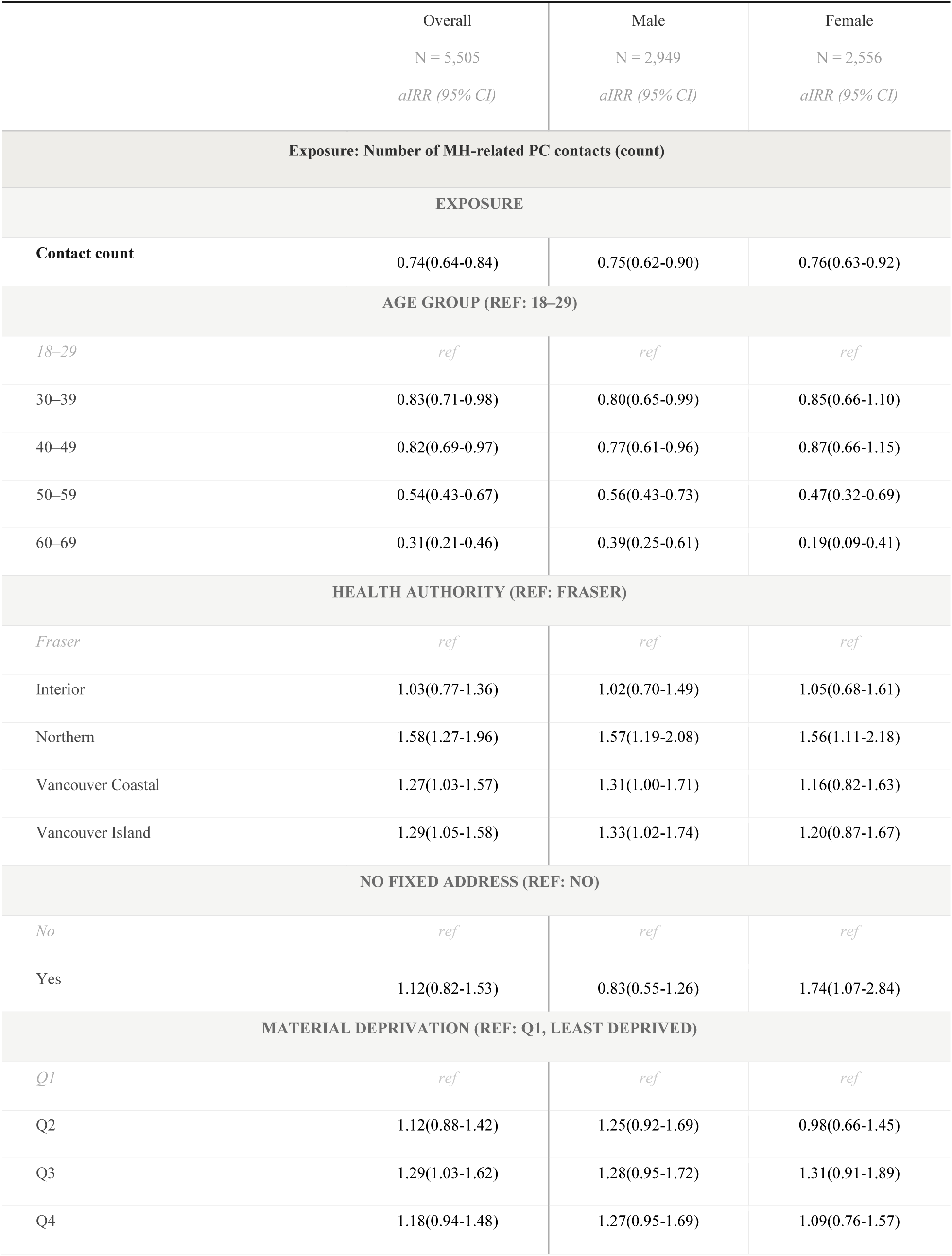
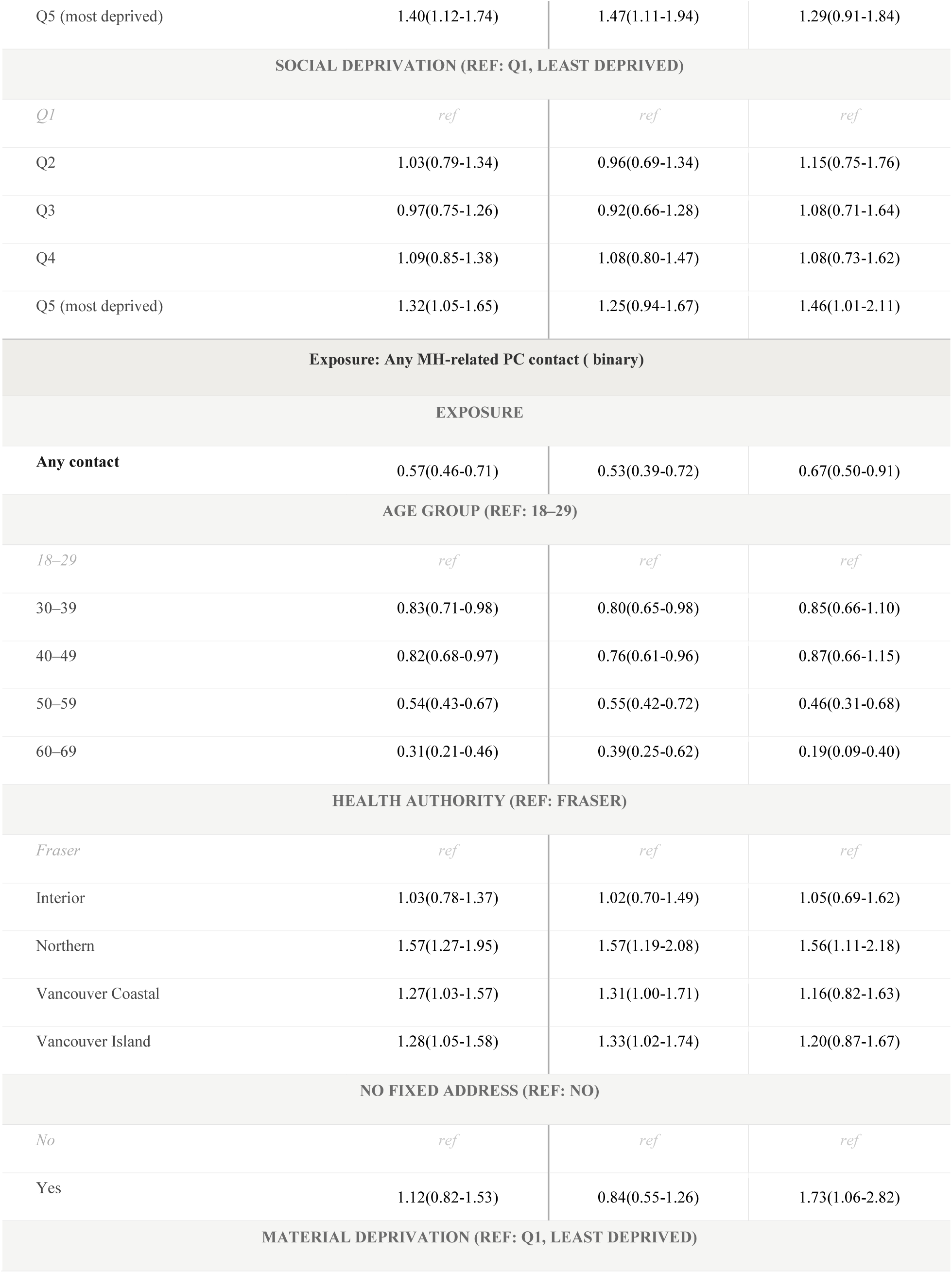
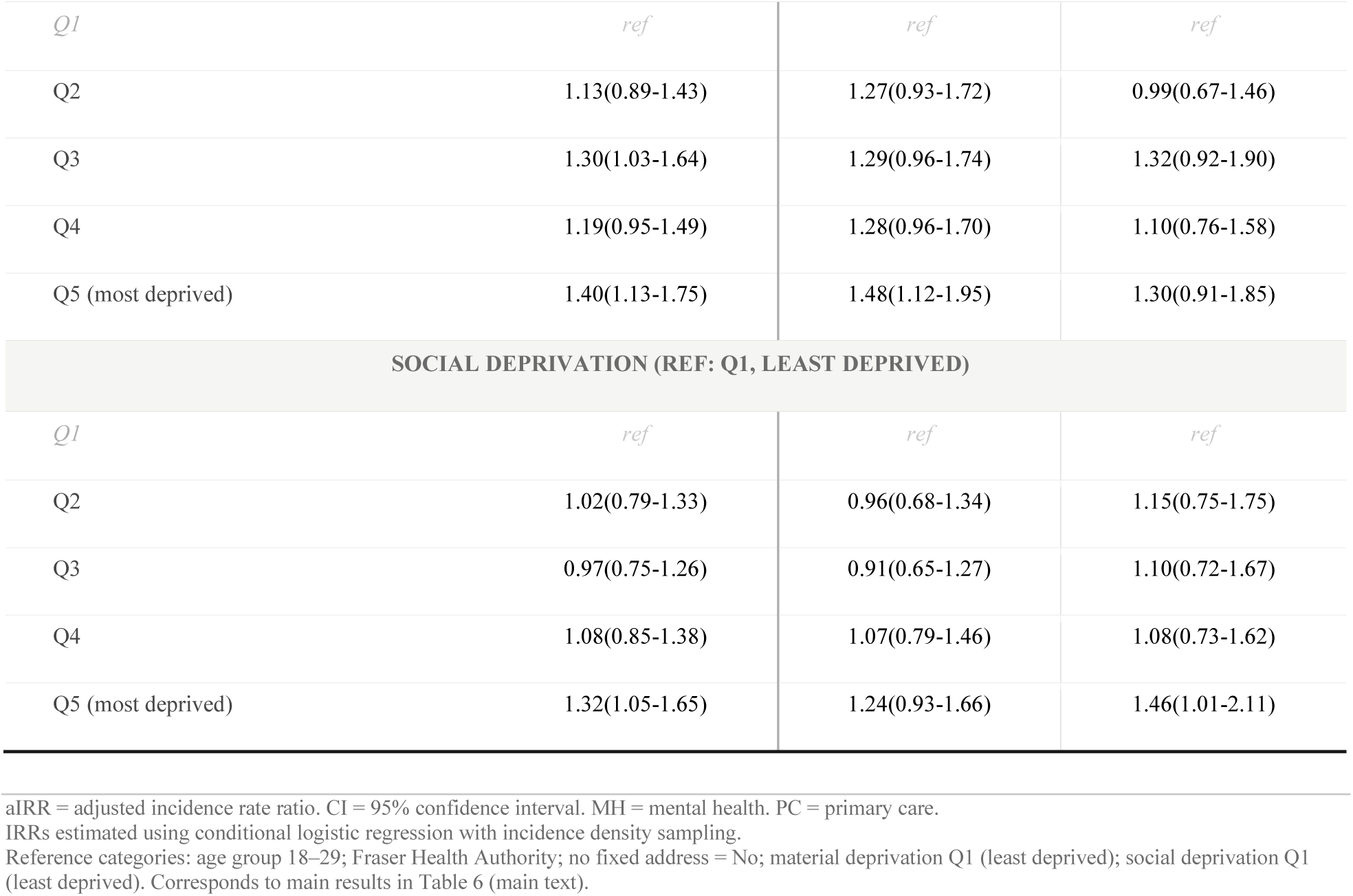
Full Model Coefficients: Association Between Mental Health-Related Primary Care Contacts and Recurrent Overdose — 30-Day Washout

**Appendix A3.** Full Model Coefficients: Association Between Mental Health-Related Primary Care Contacts and Recurrent Overdose — 90-Day Washout.

|  | Overall<br>N= 5,105<br>*aIRR (95% CI)* | Male<br>N = 2,709<br>*aIRR (95% CI)* | Female<br>N = 2,396<br>*aIRR (95% CI)* |
| --- | --- | --- | --- |
| **Exposure: Number of MH-related PC contacts (count)** |  |  |  |
| **EXPOSURE** |  |  |  |
| **Contact count** | 0.92(0.86-0.99) | 0.92(0.83-1.02) | 0.94(0.86-1.04) |
| **AGE GROUP (REF: 18–29)** |  |  |  |
| *18–29* | *ref* | *ref* | *ref* |
| 30–39 | 0.89(0.73-1.08) | 0.82(0.64-1.06) | 0.97(0.72-1.31) |
| 40–49 | 0.80(0.64-0.99) | 0.72(0.54-0.96) | 0.88(0.63-1.24) |
| 50–59 | 0.57(0.44-0.74) | 0.63(0.46-0.86) | 0.44(0.27-0.70) |
| 60–69 | 0.34(0.21-0.53) | 0.46(0.28-0.77) | 0.15(0.06-0.41) |
| **HEALTH AUTHORITY (REF: FRASER)** |  |  |  |
| *Fraser* | *ref* | *ref* | *ref* |
| Interior | 1.02(0.73-1.44) | 0.99(0.62-1.57) | 1.09(0.66-1.82) |
| Northern | 1.35(1.03-1.77) | 1.41(0.99-2.02) | 1.26(0.82-1.92) |
| Vancouver Coastal | 1.35(1.05-1.74) | 1.44(1.04-2.00) | 1.16(0.77-1.75) |
| Vancouver Island | 1.37(1.07-1.76) | 1.40(1.01-1.94) | 1.33(0.90-1.96) |
| **NO FIXED ADDRESS (REF: NO)** |  |  |  |
| *No* | *ref* | *ref* | *ref* |
| Yes | 1.20(0.83-1.74) | 0.98(0.61-1.56) | 1.64(0.89-3.01) |
| **MATERIAL DEPRIVATION (REF: Q1, LEAST DEPRIVED)** |  |  |  |
| *Q1* | *ref* | *ref* | *ref* |
| Q2 | 1.16(0.87-1.54) | 1.29(0.90-1.85) | 1.01(0.64-1.60) |
| Q3 | 1.32(1.01-1.74) | 1.31(0.92-1.87) | 1.34(0.87-2.06) |
| Q4 | 1.17(0.89-1.53) | 1.23(0.87-1.75) | 1.10(0.72-1.70) |
| Q5 (most deprived) | 1.28(0.98-1.67) | 1.37(0.98-1.92) | 1.15(0.75-1.76) |
| **SOCIAL DEPRIVATION (REF: Q1, LEAST DEPRIVED)** |  |  |  |
| *Q1* | *ref* | *ref* | *ref* |
| Q2 | 1.05(0.76-1.45) | 1.02(0.67-1.56) | 1.10(0.66-1.84) |
| Q3 | 1.04(0.75-1.43) | 1.09(0.72-1.64) | 0.99(0.59-1.64) |
| Q4 | 1.12(0.83-1.51) | 1.18(0.80-1.73) | 1.01(0.63-1.64) |
| Q5 (most deprived) | 1.40(1.06-1.85) | 1.38(0.96-1.99) | 1.45(0.93-2.24) |
| **Exposure: Any MH-related PC contact (binary)** |  |  |  |
| **EXPOSURE** |  |  |  |
| **Any contact (Yes vs No)** | 0.78(0.65-0.94) | 0.77(0.60-0.99) | 0.84(0.63-1.11) |
| **AGE GROUP (REF: 18–29)** |  |  |  |
| *18–29* | *ref* | *ref* | *ref* |
| 30–39 | 0.89(0.73-1.08) | 0.82(0.63-1.06) | 0.97(0.72-1.32) |
| 40–49 | 0.79(0.63-0.98) | 0.71(0.54-0.95) | 0.88(0.63-1.24) |
| 50–59 | 0.57(0.44-0.74) | 0.62(0.45-0.85) | 0.43(0.27-0.70) |
| 60–69 | 0.34(0.21-0.53) | 0.47(0.28-0.78) | 0.15(0.05-0.41) |
| **HEALTH AUTHORITY (REF: FRASER)** |  |  |  |
| *Fraser* | *ref* | *ref* | *ref* |
| Interior | 1.02(0.72-1.43) | 0.98(0.62-1.56) | 1.09(0.65-1.81) |
| Northern | 1.33(1.02-1.75) | 1.40(0.98-2.00) | 1.25(0.82-1.90) |
| Vancouver Coastal | 1.34(1.04-1.73) | 1.43(1.03-1.99) | 1.15(0.77-1.74) |
| Vancouver Island | 1.36(1.06-1.75) | 1.39(1.01-1.93) | 1.32(0.90-1.95) |
| **NO FIXED ADDRESS (REF: NO)** |  |  |  |
| *No* | *ref* | *ref* | *ref* |
| Yes | 1.19(0.82-1.72) | 0.97(0.61-1.54) | 1.63(0.88-2.99) |
| **MATERIAL DEPRIVATION (REF: Q1, LEAST DEPRIVED)** |  |  |  |
| *Q1* | *ref* | *ref* | *ref* |
| Q2 | 1.16(0.87-1.53) | 1.29(0.90-1.85) | 1.01(0.64-1.61) |
| Q3 | 1.32(1.00-1.73) | 1.30(0.91-1.85) | 1.34(0.87-2.07) |
| Q4 | 1.17(0.89-1.53) | 1.23(0.87-1.74) | 1.10(0.72-1.70) |
| Q5 (most deprived) | 1.28(0.98-1.66) | 1.36(0.97-1.91) | 1.15(0.75-1.76) |
| **SOCIAL DEPRIVATION (REF: Q1, LEAST DEPRIVED)** |  |  |  |
| *Q1* | *ref* | *ref* | *ref* |
| Q2 | 1.05(0.75-1.45) | 1.02(0.67-1.56) | 1.10(0.66-1.83) |
| Q3 | 1.04(0.75-1.43) | 1.08(0.72-1.63) | 0.99(0.60-1.65) |
| Q4 | 1.12(0.83-1.51) | 1.18(0.80-1.73) | 1.01(0.63-1.64) |
| Q5 (most deprived) | 1.40(1.05-1.85) | 1.38(0.96-1.99) | 1.45(0.93-2.24) |

aIRR = adjusted incidence rate ratio. CI = 95% confidence interval. MH = mental health. PC = primary care.
IRRs estimated using conditional logistic regression with incidence density sampling.

**Appendix A4.** Sensitivity Analysis: Association Between Mental Health-Related Primary Care Contacts and Recurrent Overdose Among People with a History of a Depression Diagnosis Using Unconditional Logistic Regression with Natural cubic Spline Adjustment for Person-Time at Risk.

|  | aOR (95% CI) | p |
| --- | --- | --- |
| **30-day washout** |  |  |
| OVERALL (N = 5,505) |  |  |
| *Contact count* | 0.73(0.64-0.84) | p<0.001 |
| *Any contact (binary)* | 0.58(0.46-0.72) | p<0.001 |
| MALE (N= 2,949) |  |  |
| *Contact count* | 0.75(0.62-0.91) | p<0.05 |
| *Any contact (binary)* | 0.53(0.39-0.73) | p<0.001 |
| FEMALE (N= 2,556) |  |  |
| *Contact count* | 0.75(0.62-0.91) | p<0.01 |
| *Any contact (binary)* | 0.66(0.49-0.90) | p<0.01 |
| **90-day washout** |  |  |
| OVERALL (N = 5,105) |  |  |
| *Contact count* | 0.92(0.86-0.99) | p<0.05 |
| *Any contact (binary)* | 0.78(0.65-0.94) | p<0.01 |
| MALE (N = 2,709) |  |  |
| *Contact count* | 0.92(0.83-1.02) | n.s. |
| *Any contact (binary)* | 0.77(0.59-0.99) | p<0.05 |
| FEMALE (N= 2,396) |  |  |
| *Contact count* | 0.95(0.86-1.05) | n.s. |
| *Any contact (binary)* | 0.85(0.64-1.13) | n.s. |

aOR = adjusted odds ratio; CI = 95% confidence interval. Contact count refers to the total number of mental health-related primary care contacts within the specified exposure window. Any contact refers to the presence of at least one mental health-related primary care contact within the specified exposure window. Person-time at risk was included as a covariate modelled using a natural cubic spline.

